# Genetic Risk of Reticular Pseudodrusen in Age-Related Macular Degeneration: *HTRA1*/lncRNA *BX842242.1* dominates, with no evidence for Complement Cascade involvement

**DOI:** 10.1101/2024.09.26.24314339

**Authors:** Samaneh Farashi, Carla J Abbott, Brendan RE Ansell, Zhichao Wu, Lebriz Altay, Ella Arnon, Louis Arnould, Yelena Bagdasarova, Konstantinos Balaskas, Fred K Chen, Emily Chew, Itay Chowers, Steven Clarke, Catherine Cukras, Cécile Delcourt, Marie-Noëlle Delyfer, Anneke I. den Hollander, Sascha Fauser, Robert P. Finger, Pierre-Henry Gabrielle, Jiru Han, Lauren AB Hodgson, Ruth Hogg, Frank G Holz, Carel Hoyng, Himeesh Kumar, Eleonora M Lad, Aaron Lee, Ulrich FO Luhmann, Matthias M Mauschitz, Amy J McKnight, Samuel McLenachan, Aniket Mishra, Ismail Moghul, Luz D Orozco, Danuta M Sampson, Liam W Scott, Vasilena Sitnilska, Scott Song, Amy Stockwell, Anand Swaroop, Jan H Terheyden, Liran Tiosano, Adnan Tufail, Brian L Yaspan, MACUSTAR consortium, NICOLA consortium, Alice Pébay, Erica L Fletcher, Robyn H Guymer, Melanie Bahlo the Reticular Pseudodrusen Consortium

## Abstract

Age-related macular degeneration (AMD) is a multifactorial retinal disease with a large genetic risk contribution. Reticular pseudodrusen (RPD) is a sub-phenotype of AMD with a high risk of progression to late vision threatening AMD. In a genome-wide association study of 2,165 AMD+/RPD+ and 4,181 AMD+/RPD-compared to 7,660 control participants, both chromosomes 1 (*CFH*) and 10 (*ARMS2/HTRA1*) major AMD risk loci were reidentified. However association was only detected for the chromosome 10 locus when comparing AMD+/RPD+ to AMD+/RPD-cases. The chromosome 1 locus was notably absent. The chromosome 10 RPD risk region contains a long non-coding RNA (ENSG00000285955/BX842242.1) which colocalizes with genetic markers of retinal thickness. *BX842242.1* has a strong retinal eQTL signal, pinpointing the parafoveal photoreceptor outer segment layer. Whole genome sequencing of phenotypically extreme RPD cases identified even stronger enrichment for the chromosome 10 risk genotype.

## Introduction

Age-related macular degeneration (AMD) is a leading cause of severe irreversible vision loss worldwide (1). Late-stage AMD complications result from damage to the photoreceptors and their supporting cells, the retinal pigment epithelium (RPE) (2,3).

The hallmark of AMD is the presence of drusen, which are focal accumulations of extracellular, lipid-rich debris underneath the RPE. Their increasing size and extent are associated with an increased risk of progression to vision-threatening late AMD (4). Reticular pseudodrusen (RPD), or subretinal drusenoid deposits (SDD), refer to distinct deposits that accumulate apical to the RPE (or in the subretinal space) (5). They are present in approximately 30% of individuals with early stage AMD, and up to 60% of those with late-stage disease. RPD are a critical AMD sub-phenotype driving vision loss (5) and have been associated with a higher risk of AMD progression, worse visual function, and poorer treatment outcomes (6).

AMD is a complex multifactorial disease, with a strong genetic predisposition, and an incompletely understood pathophysiology. Similarly, the pathogenesis of RPD and the biologic basis for their contribution to poor prognosis and visual outcomes is also not understood. Several large GWAS studies have identified genetic loci at the complement factor H (*CFH*) gene family at the *1q31* locus and the *ARMS2/HTRA1* genes at the *10q26* locus, as having the largest effects for AMD risk (7,8). In contrast, previous genetic studies in RPD have not performed GWAS analysis, but rather have limited their analysis to selected AMD-associated SNPs within the *ARMS2/HTRA1, C3* and *VEGFA* genes (9).

We conducted the first genome-wide association study (GWAS) to identify the genetic risk factors associated specifically with RPD. Joint analysis with outer retinal traits and retinal expression data has leveraged new insights from genome annotations and confirmed results with genome sequencing and RPD load analyses (Fig. 1). This study sheds light on genetic risk factors for RPD and how these function in the retina, providing a foundational understanding for future investigations.

**Fig. 1:**
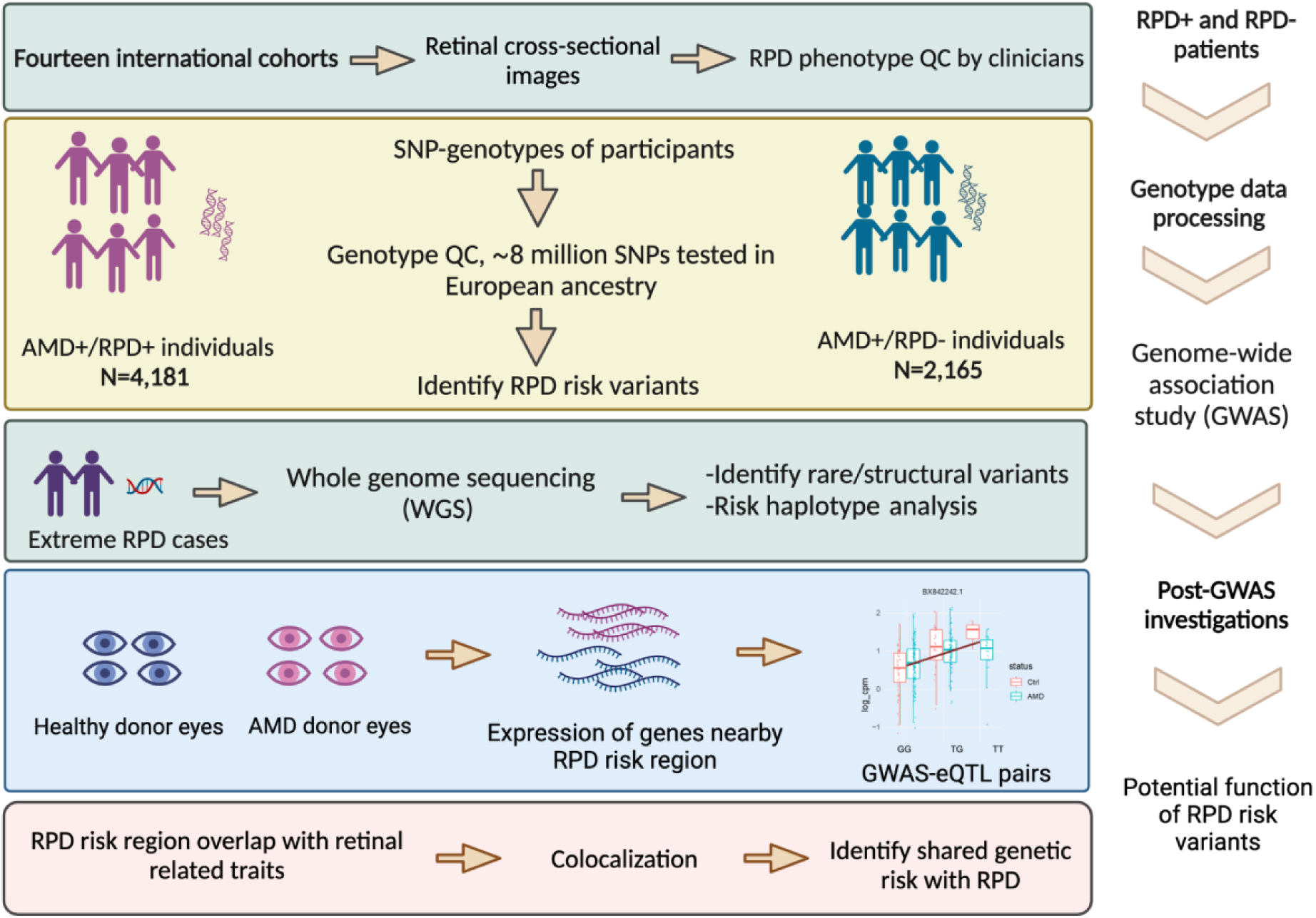
Overall study design in multiple stepwise strategies from cohort collection, GWAS to subsequent post-GWAS analyses.

## Results

### Harmonization of phenotyping and genotyping

Fourteen cohorts contributed to the RPD consortium, where RPD were primarily defined based upon their presence in optical coherence tomography (OCT) imaging, the optimal method to detect RPD (Supplementary Table S1). Ten cohorts contributed individual-level genotype data and the remaining four cohorts shared summary statistics results of GWAS (Supplementary Table S2).

Phenotyping harmonization of the individual-level data led to a dataset of 3,109 AMD+/RPD-, 1,532 AMD+/RPD+ and 2,083 controls (AMD-/RPD-) (Supplementary Table S3).

Genotyping data was harmonized by combining cohorts by origin, e.g. whole genome sequencing, or SNP array platform (Supplementary Table S4). Genotyping quality control (QC) processes led to the identification of a set of high-quality variants across cohorts to facilitate the meta-analysis (Supplementary Table S5). Curation of the genotype data after QC resulted in a dataset of 2,165 AMD+/RPD+; 4,181 AMD+/RPD-, and 7,660 AMD-/RPD-controls (total samples 14,006).

Detailed demographic data are shown in Supplementary Table S3 and Supplementary Fig. S1. Recruitment strategies influenced the range of AMD and RPD prevalence across cohorts (Supplementary Fig. S2).

### GWAS meta-analysis

We first conducted an AMD GWAS with available individual-level data in AMD+(RPD+/-) (N=3,959) versus AMD-/RPD-controls (N=1,946), serving as a QC step to confirm sensitivity and power to detect known key AMD risk loci (Fig. 2a). In this overall AMD-GWAS, we detected genome-wide significant association signals on chromosome 1, 6 and 10 confirming the three major AMD risk loci identified previously (Supplementary Table S6) (7,10).

**Fig. 2:**
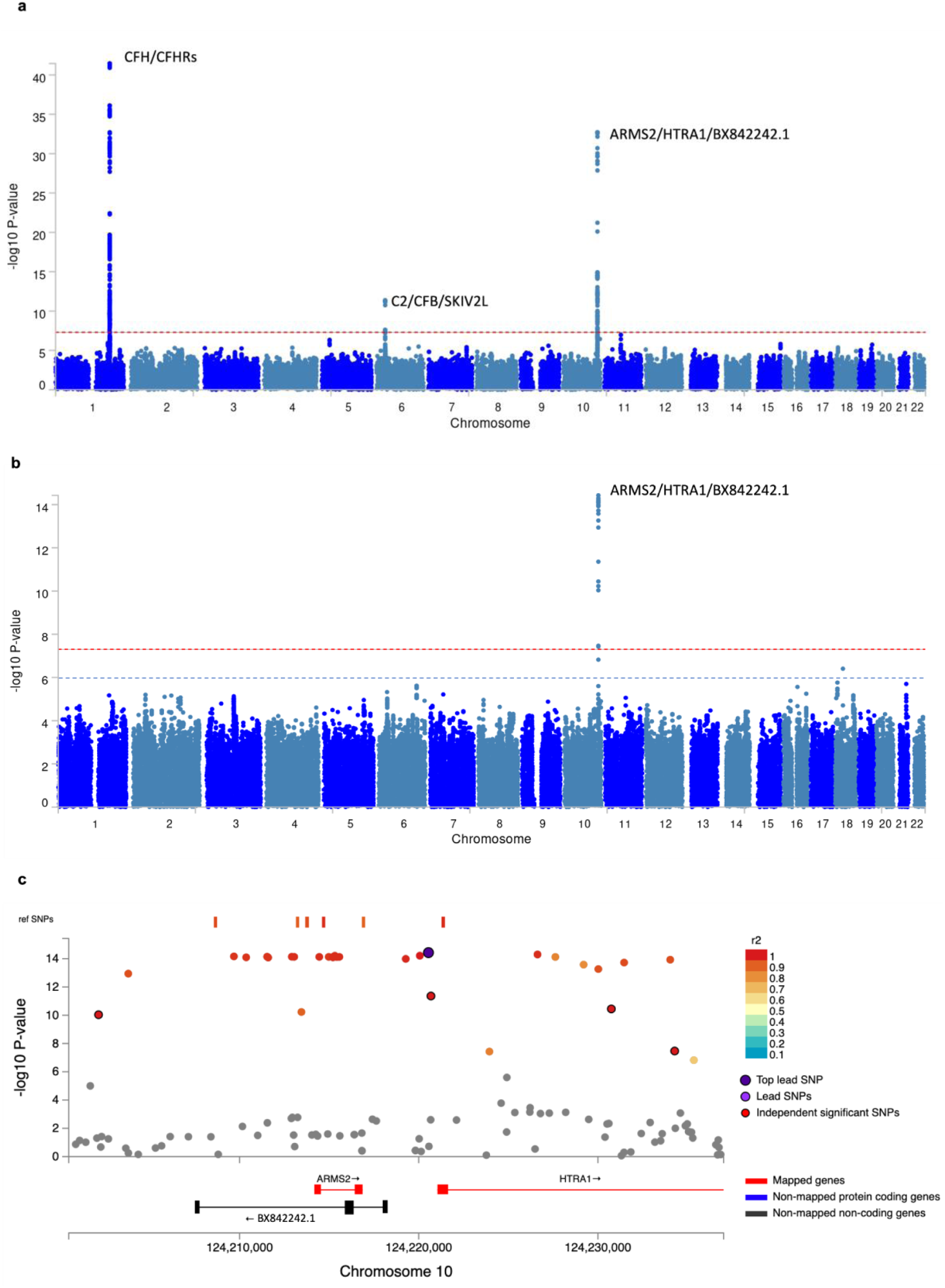
Results of Genome-wide association studies (GWAS). **a** Manhattan plot of overall AMD-GWAS meta-analysis showing –log10(*P-value*) values across the genome comparing AMD+ (RPD+/-) participants (N= 3,959) vs. AMD-controls (N= 1,946). The red horizontal line represents the genome-wide significance level (*P-value* = 5 × 10^-8^). Nearby genes to the two main associated loci with AMD risk on chromosomes 1, 6 and 10 are labelled; **b** Manhattan plot of the meta-analysis of AMD+/RPD+participants compared to AMD+/RPD-participants. Common variants display a genome-wide significant locus on chromosome 10. The y-axis indicates −log10(*P-value*). The red and blue horizontal lines represent the genome-wide significance level (*P-value* = 5 × 10^-8^) and suggestive genome-wide significance level (*P-value* = 1 × 10^-6^), respectively. Note that the RPD-specific GWAS found only the *ARMS2/HTRA1* risk locus on chromosome 10 and the complement cascade risk factors on chromosome 1 (*CFH/CFHRs*) were absent; **c** Regional plot with annotations displaying the top RPD-associated signal. The color of each point (SNP) is determined by its linkage disequilibrium (LD) squared correlation coefficient (r^2^) with respect to the top lead variant, displayed in dark purple (rs11200638). Independent significant SNPs are displayed as red circles outlined in black. Genes proximal to this RPD risk locus are labeled, including *ARMS2*, *HTRA1* (both in red) and the long non-coding RNA *BX842242.1* (in black). *BX842242.1* was added manually. The orange dashes over the top of the SNPs indicate SNPs in the reference genome (1000G Phase 3 European ancestry). Genomic locations are plotted using the hg19 genomic build.

We then performed an RPD-specific GWAS meta-analysis comparing AMD+/RPD+ cases to AMD+/RPD-cases, identifying the genetic risk loci specific for RPD. Prior to the meta-analysis, genomic control correction (GC) was applied to each GWAS to correct for population stratification. The GC inflation factor (λ) ranged from 0.9 to 1.3 (Supplementary Table S5). The veracity of results for individual GWAS was confirmed using Quantile-Quantile plots (Q-Q plots, Supplementary Fig. S3a).

After frequency annotation and quality control steps, the number of SNPs varied between cohorts, with a minimum of 6,984,307 available SNPs for analysis to a maximum of 8,369,653 SNPs (with minor allele frequency (MAF) of > 0.01), for inclusion in the meta-analysis (Supplementary Table S5). A GWAS meta-analysis in European populations across the 14 cohorts for the 2,165 AMD+/RPD+ and 4,181 AMD+/RPD-samples was performed using an inverse-variance weighted fixed effect meta-analysis method, implemented via METAL (11). The GC λ of the GWAS meta-analysis summary statistics was 1.01 (Supplementary Fig. S3b).

The GWAS identified one risk region on chromosome 10 at the ARMS2/HTRA1 locus (rs11200638, beta = 0.26, p=3.73e^-15^), reaching genome-wide significance (*P-value* < 5 × 10^−8^) (Fig. 2b). Two protein-coding genes, *ARMS2* and *HTRA1* are located at this risk locus (Fig. 2c), with genome-wide significantly associated SNPs spanning both genes. In addition to *ARMS2* and *HTRA1,* a *lncRNA* gene (*BX842242.1/ENSG00000285955*; Fig. 2c) is located within the RPD risk region.

One locus on chromosome 18 (SNP rs76361335) was identified as associated with RPD at suggestive significance (*P-value* < 1 × 10^−6^) (Supplementary Table S7). However, this SNP has low confidence due to its low MAF (0.02 and 0.011 in cases and controls, respectively) and because it is an orphan signal with no other associated SNPs in LD. The SNP resides in a gene desert (Supplementary Fig. S4).

The genome-wide significant region identified in the RPD GWAS contains multiple SNPs in high LD (Supplementary Fig. S5a), but conditional analysis on the RPD-risk lead SNP (rs11200638) did not reveal any additional independent SNPs (Supplementary Fig. S5b).

Our primary GWAS did not identify any significant variants within the canonical ‘Complement factor related genes’ risk loci on chromosome 1 in AMD, the most prominent signal in previous AMD GWAS (10). Power studies suggested 100% power to detect the top significant SNP on chromosome 1 (rs800292) from a recent AMD-GWAS (10), with beta=-0.7 and MAF=0.23, in our RPD-GWAS at the genome-wide significance level (*P-value* < 5 × 10^−8^) (Supplementary Fig. S6).

Two additional GWAS were performed: (i) AMD+/RPD+ versus controls (N=1,852 and N=7,660, respectively: ‘GWAS2’), and (ii) AMD+/RPD-versus controls (N=3,607 and N=7,660, respectively: ‘GWAS3’; Supplementary Fig. S7a). Two distinct meta-analyses were carried out by incorporating the summary statistics data obtained from GWAS2 and GWAS3 for the eight cohorts with feasible control samples (Supplementary Table S8).

Genome-wide significant associations with the CFH gene family on chromosome 1 were identified in both ancillary studies with similar effect sizes to those previously described for AMD (Supplementary Table S9 & Supplementary Fig. S7b:e) while the risk loci on chromosome 1 were notably absent in the RPD specific GWAS (‘GWAS1’) (Fig. 2a:b).

To further investigate the GWAS finding on chromosome 10, we performed colocalization analyses across GWAS1, GWAS2 and GWAS3 and compared it to the results of two published AMD GWAS by Fritsche *et al* and Han *et al* (7,10). Colocalization analyses with the risk variants within the *ARMS2/HTRA1* locus confirmed that the identified region corresponds to the same locus previously identified in AMD GWAS (Supplementary Fig. S8). Notably, there was complete colocalization (posterior probability of 1.0) between all pairs of GWAS 2, 3 and GWAS results from Fritsche et al., and Han et al., with a 90% overlap (posterior probability of 0.90) observed in GWAS1 variants compared to the other four GWAS studies (Supplementary Fig. S8).

X chromosome analysis, feasible for a subset of cohorts (eight cohorts) (AMD+/RPD+: N=1,707, AMD+/RPD-: N=3,321; Supplementary Table S8), identified no significant signals.

### Study-specific association analyses of RPD risk region

Study-specific logistic regression association analyses were performed using an additive genotypic association model following a predetermined/RPD study-specific analysis plan. We examined the RPD-risk lead SNP (rs11200638) in each summary statistic of individual cohorts to explore its significance and effect size (Supplementary Fig. S9a).

To assess the influence of each cohort on the meta-analysis, we systematically excluded one cohort at a time, conducted separate meta-analyses, and compared the *P-values* and effect sizes of the RPD risk lead SNP across these analyses. The lead SNP remained genome-wide significant in all leave-one-out cohort permutations (Supplementary Fig. S9b).

### Association between RPD lesion extent and RPD risk region

An AI algorithm applied to the OCT imaging data quantified the macular extent of RPD in a subset of AMD+/RPD+ individuals (12). We investigated the association between the chromosome 10 RPD risk locus and the extent of RPD while adjusting for age and sex. The Findings reveal the RPD risk lead SNP (rs11200638) is significantly associated with RPD extent, with more pronounced effects observed in higher RPD extent deciles (Fig. 3a). In contrast, the chromosome 1 risk SNP (rs10922109; *CFH*) is not associated with RPD extent (Fig. 3b). A strong and consistent dose dependent effect is observed between the RPD extent deciles and the RPD risk allele dosage (Fig. 3c), with a similar increase in the allele frequency of RPD risk lead SNP (Fig. 3d).

**Fig. 3.**
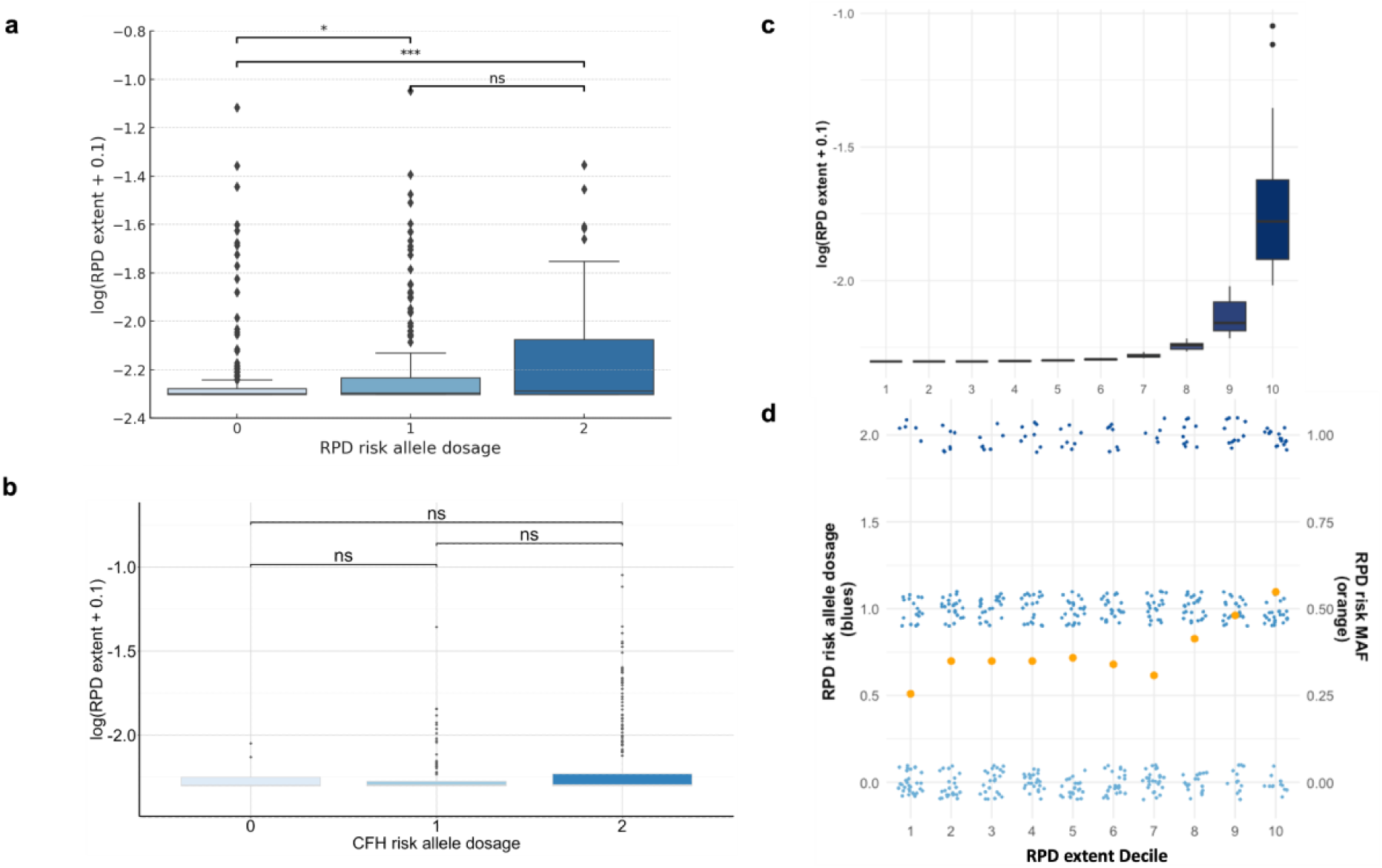
Reticular pseudodrusen (RPD) extent is significantly associated with heightened RPD genetic risk. **a** Box plot showing the RPD extent in N=526 individuals relative to the RPD-risk lead SNP (rs11200638) dosage (x-axis) and **b** Box plot showing RPD extent in N=526 individuals relative to the CFH top risk SNP (rs10922109) based on Fritsche *et al* (*7*) dosage (x-axis) while adjusting for age and sex as covariates. The y-axis represents the log-transformed RPD extent based on a percentage of the entire scan with RPD (the mean RPD extent of the left and right eyes were used) adding a constant of 0.1 to avoid log (0) of N=181 individuals who showed a mean extent RPD value of zero. A linear regression test was conducted to examine the differences in the RPD extent across the genotype groups controlling for age and sex. We performed pairwise Wilcoxon tests with Bonferroni correction to directly compare the p-values among groups. Significant differences between groups are indicated by asterisks (p > 0.05: ’ns’, p < 0.05: ’*’, p < 0.01: ’**’, p < 0.001: ’***’); **c** RPD extent as defined in panel a (y-axis), binned into deciles (x-axis) for analysis of RPD risk. **d** RPD risk allele frequency (left y-axis) relative to RPD extent deciles (x-axis); In d, blue points represent individual study participants’ genetic risk, colored by risk allele dosage. Orange points represent risk allele frequency per decile (right y-axis). The plot illustrates how the RPD-risk lead SNP genotypes vary across different levels of RPD extent, providing insights into the genetic association between RPD risk and the severity of RPD.

### Non-additive GWAS

Non-additive genetic associations have been previously identified in AMD for several risk loci in genes such as *THUMPD2* (13). To investigate the association of common variants in a non-additive manner on RPD, we conducted association analyses on the autosomes only, using both dominant and recessive models comparing AMD+/RPD+ to AMD+/RPD-(GWAS 1 in Supplementary Fig. S7a). Two meta-analyses incorporated the summary statistics from dominant and recessive models performed for seven cohorts with individual-level genotyping data for AMD+/RPD+ (N=2,165) and AMD+/RPD-cases (N=4,181) (Supplementary Table S8). We identified no genome-wide significant signal (Supplementary Fig. S10 a:d). However, SNP 10:124227624:C:T (rs60401382) within the *ARMS2/HTRA1* risk locus in the additive model was associated with RPD at the suggestive significance level under both dominant (beta= 0.23, p=2e^-07^) and recessive models (beta= 0.21, p=9e^-07^), (Supplementary Table S10) with the same effect sizes, suggesting the rs60401382 signal is underpinned by a dominant effect.

### Whole Genome Sequencing of extreme RPD phenotype participants

To further investigate the RPD risk haplotype and examine the clinical importance of the identified risk region, short-read whole genome sequencing (WGS) was conducted in a distinct AMD+ cohort (from the Centre for Eye Research Australia, CERA) of European ancestry and unrelated individuals with extreme RPD (‘AMD+/RPD++’), defined as having extensive and predominantly RPD deposits. Importantly, the extreme RPD phenotype participants were chosen masked to their RPD genetic risk profile, particularly of the chr10 risk genotype status.

The association between the extreme RPD status and the RPD risk lead SNP (rs11200638) was evaluated using logistic regression models adjusted for age, sex, and two ancestry PCs, in AMD-controls (N=1,008) compared to each of three cohorts: (1) AMD+/RPD++ (N=44), (2) AMD+/RPD+ (N=1,055), and (3) AMD+/RPD-(N=1,792). This revealed the RPD risk lead SNP was significantly associated with RPD in all three comparisons with the highest odds ratio (OR) observed in AMD+/RPD++ cases (OR=6.54, CI=(11.4 -33.76)) (Table 1). The Wald tests showed significant differences in the odds ratios between all analysis pairs (Supplementary Table S11).

**Table 1.**
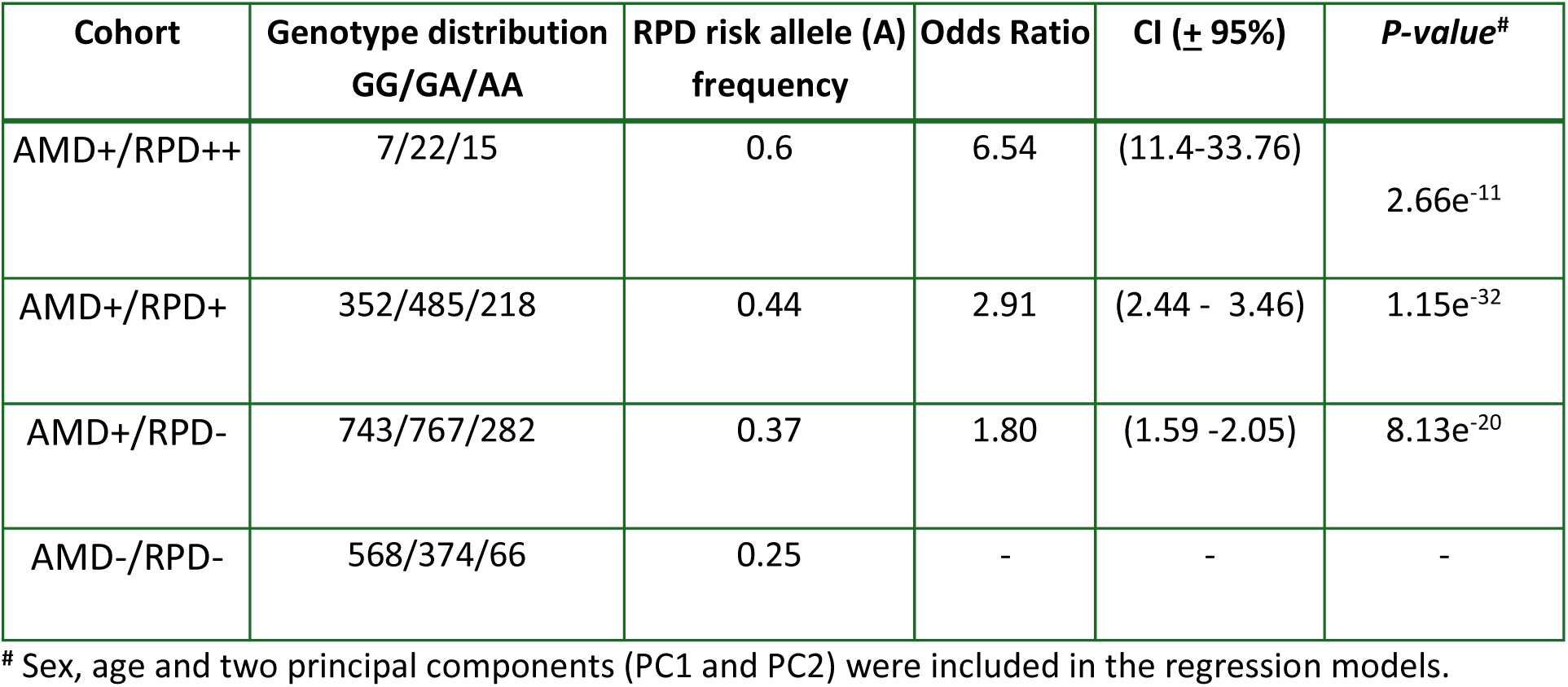
Results of logistic regression analysis of RPD risk lead SNP (rs11200638) genotypes in three cohorts compared to AMD-/RPD-controls.

| Cohort | Genotype distribution<br>GG/GA/AA | RPD risk allele (A)<br>frequency | Odds Ratio | CI (± 95%) | P-value<sup>#</sup> |
| --- | --- | --- | --- | --- | --- |
| AMD+/RPD++ | 7/22/15 | 0.6 | 6.54 | (11.4-33.76) | 2.66e<sup>-11</sup> |
| AMD+/RPD+ | 352/485/218 | 0.44 | 2.91 | (2.44 - 3.46) | 1.15e<sup>-32</sup> |
| AMD+/RPD- | 743/767/282 | 0.37 | 1.80 | (1.59 -2.05) | 8.13e<sup>-20</sup> |
| AMD-/RPD- | 568/374/66 | 0.25 | - | - | - |

<sup>#</sup> Sex, age and two principal components (PC1 and PC2) were included in the regression models.

The RPD risk lead SNP (rs11200638) within the *ARMS2/HTRA1* risk locus was found to have the highest frequency (0.6) in AMD+/RPD++. The odds ratio of the RPD risk effect allele (A allele) of SNP rs11200638 in AMD+/RPD++ is more than double that of AMD+/RPD+ (Table 1). The MAF of the risk lead SNP was observed to be ∼3-fold higher in AMD+/RPD++ (MAF=0.6) compared to non-AMD controls (MAF=0.25) or non-Finnish Europeans in gnomAD v3.2.1 (MAF=0.22) (14).

We investigated the WGS data in AMD+/RPD++ individuals to identify potentially pathogenic rare genetic variants (MAF > 0.001) and structural variants in the genes within the RPD risk locus (*ARMS2* and *HTRA1*) including coding, intronic and untranslated regions (UTRs) using a rare-variant GATK pipeline analysis. After filtering with gnomAD v3.2.1 (14) and other QC checks, no variants predicted to be deleterious remained. Using the WGS data, we identified 198 other variants within the RPD-risk region genomic window (chr10:124205544- 124235544), of which 83 variants existed in the 1000 Genome reference panel allowing LD testing with the RPD-risk lead SNP. LD scores ranged from r^2^=0.004 (multiple SNPs) to r^2^=1 (rs3763764) with multiple LD blocks (Supplementary Fig. S11).

The WGS-based structural variation analysis led to the identification of a known insertion/deletion polymorphism (del443ins54) in the 3′UTR of *ARMS2* and within the intronic region of lncRNA *BX842242.1* with a MAF of 0.31 in the non-Finnish European population (based on gnomAD SVs v4.3.1). This indel is in strong LD with the lead SNP (rs11200638) (r^2^=0.98 in the Australian population, Supplementary Fig. S12a). Another coding variant, rs10490924 (c.205G>T, p.Ala69Ser), with a high *in-silico* score for predicted damaging variants (CADD score:15.87) was in LD with the risk haplotype (Supplementary Fig. S12b).

### RPD risk haplotype characterization

The WGS data in individuals homozygous for the chromosome 10 risk tagging haplotype (rs11200638; N=15) for the RPD risk lead variant was used to perform haplotype analysis of the RPD risk locus. Critical region analysis identified a 95% confidence region spanning rs1591008260 (chr 10:124197751) to rs1591024450 (chr 10:124227605-124227614) of total length ∼30 kb. Analysis of this region in individuals homozygous for the chromosome 10 risk tagging haplotype (rs11200638) revealed five haplotype blocks with spans ranging in size from 55 bp to 7.64 kb (Supplementary Table S12).

### Consistent induction of a novel lncRNA and suppression of *HTRA1 inferred in RPD retina*

The lncRNA *BX842242.1* was first included in GENCODE release 29 (late 2018). To correct for annotation variation based on the usage of different human genome references in different retinal data sets for homogenous retinal eQTL curation, we re-mapped two independent human post-mortem retina RNAseq datasets to transcriptionally quantify this alongside *ARMS2*, *HTRA1* and *PLEKHA1*. *PLEKHA1* has been identified as a risk gene for AMD, lies upstream of the key risk region associated with RPD and is included within the critical 10q26 region associated with AMD susceptibility. The ‘NEI cohort’ included matched retinal RNAseq and genotype data for 404 donors (15,16). The ‘Genentech cohort’ included up to four tissues (neural retina and RPE; macula and non-macula) from 119 donors (15,16).

ARMS2 did not survive expression thresholding in either cohort. eQTL analysis revealed a significant positive correlation between the rs11200638 risk allele (A) dose and abundance of BX842242.1 in the neural retina (non-macula) in both cohorts (p < 10^-11^, adj.p = 0.02 respectively). Positive associations in the neural retina (macula) and RPE (non-macula) were also detected (adj.p = 0.02; Genentech) (Supplementary Table S13). Negative associations with HTRA1 were detected in the neural retina (NEI; p = 0.002), neural macula (adj.p = 0.02; Genentech), and macula and non-macula RPE regions (adj.p = 0.04 & 0.03 respectively) (Supplementary Table S14) (Fig. 4). Despite the consistent QTL results, the correlation in RNA expression between HTRA1 and BX842242.1 varied substantially across tissues and studies (e.g. non-macula retina expression was positively correlated in the NEI cohort but negatively correlated in the Genentech cohort (Supplementary Fig. S13)).

**Fig. 4:**
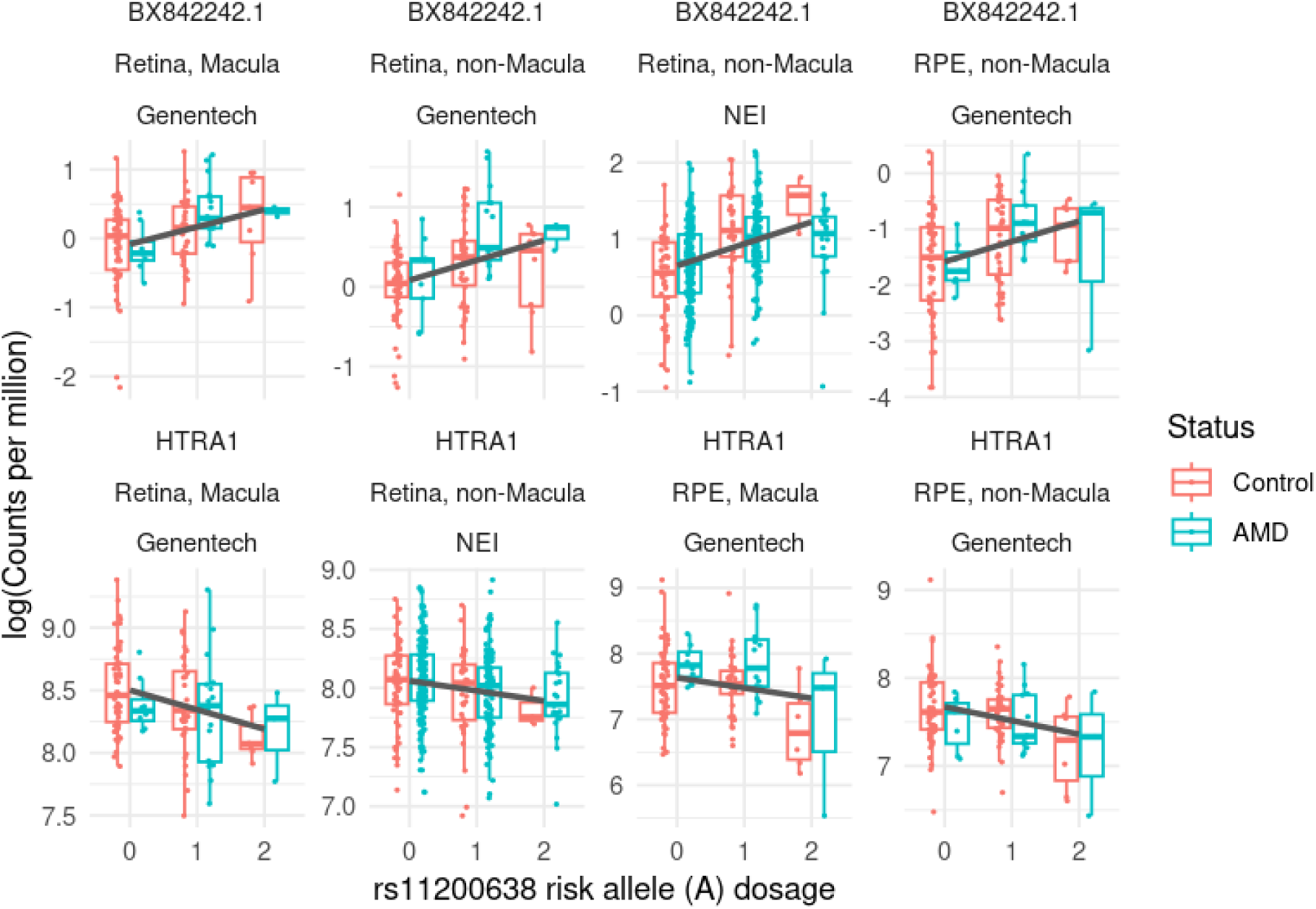
Increased RPD genetic risk is associated with increased expression of *BX842242.1* retinal RNA, and decreased expression of *HTRA1*. Only significant eQTL results are displayed. *BX842242.1*: long non-coding RNA gene; *HTRA1*: HtrA serine peptidase 1. The y-axis represents the log-transformed expression level of each gene; the x-axis denotes the risk allele dosage of the RPD risk lead variant (0: individuals who carry no RPD risk allele, 1: heterozygous for RPD risk allele and 2: homozygous for RPD risk allele). Regression testing corrected for disease status (single regression line).

Colocalization of published eQTL summary statistics for BX842242.1 (17) overlapped with 26 out of 29 genome-wide significant RPD risk-associated SNPs (Supplementary Fig. S14). Colocalization indicated a shared haplotype underlying RPD risk, and differential expression of BX842242.1 (posterior probability of colocalization [PPH4] = 0.97) (Fig. 5).

**Fig. 5.**
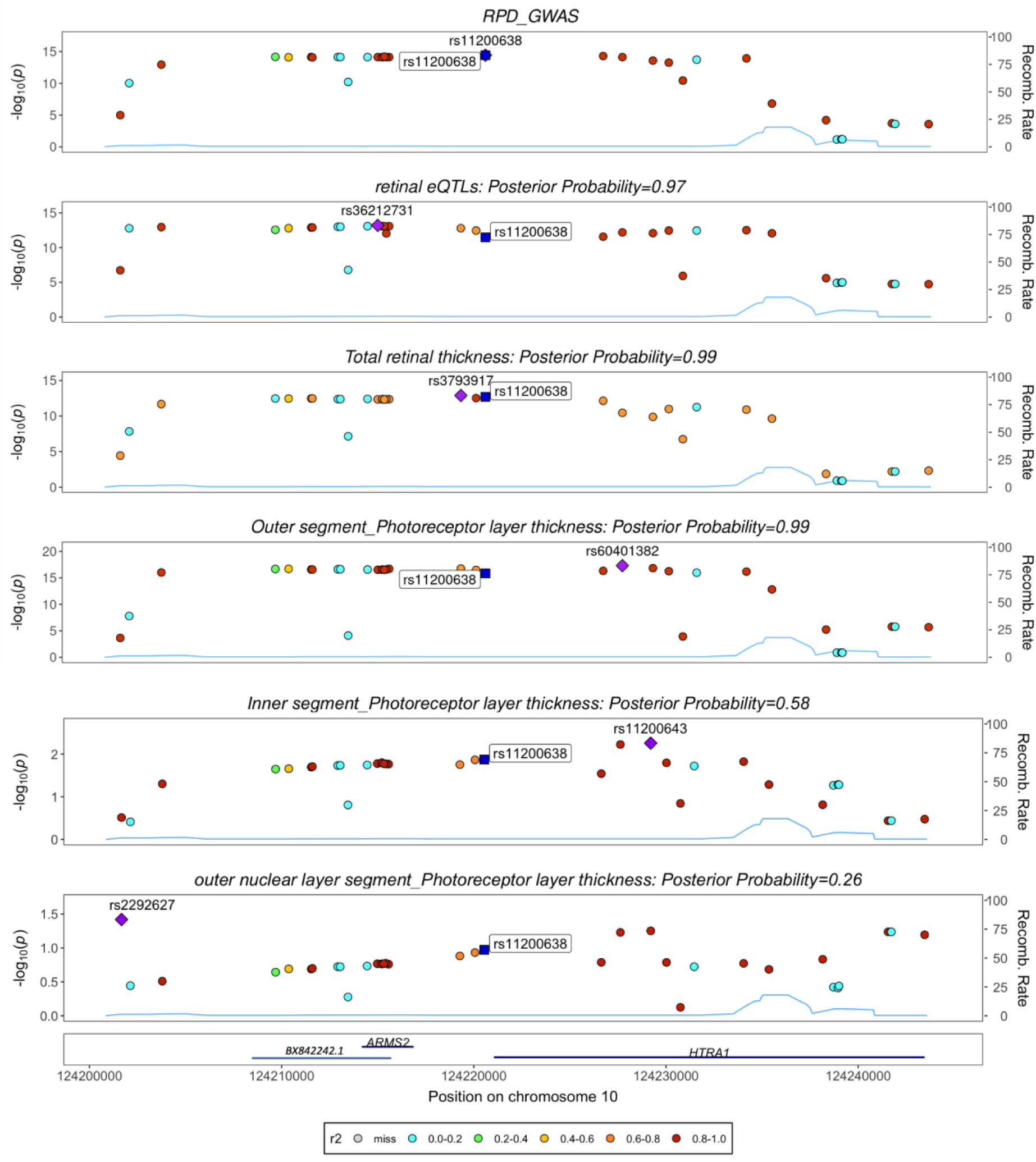
Genome-wide multi-trait colocalization analysis of RPD and candidate traits that show *ARMS2/HTRA1* locus as a significant risk-associated region. Stacked association plots of traits with > 95% shared variants with RPD. Colocalization analysis with Coloc implicates the RPD risk lead variant rs11200638 as being the shared causal variant between RPD risk and eQTLs, photoreceptor cell outer segment (OS) thickness and total retinal thickness. In contrast, the thickness of photoreceptor cell layers including the inner segment (IS) and photoreceptor cell outer nuclear layer (ONL) did not show a high overlap with the RPD risk region. Each plot includes a recombination rate track (blue line) and a linkage disequilibrium (LD) color scale indicating the r² values of SNPs in relation to RPD-risk lead SNP (rs11200638). The x-axis represents the position on chromosome 10, and the y-axis represents the -log10(*P-value*) of association. The blue bars on the x-axis label the genes. lncRNA *BX842242.1* was added manually. The purple diamond represents the most significant SNP in the region for each trait and the dark blue square represents the RPD-risk lead SNP (rs11200638) in each trait.

### RPD risk region and cross-trait analysis

Six retinal related traits with available summary statistics results, including total retinal thickness, the thickness of photoreceptor cell inner segments (IS), photoreceptor cell outer nuclear layer (ONL), photoreceptor cell outer segments (OS), retinal editing quantitative trait loci (edQTLs) and retinal methylation quantitative trait loci (mQTL) were investigated for association with the chromosome 10 RPD risk locus. The GWAS results from total retinal thickness (including RPE) and sub-cellular level of photoreceptors were performed using the UK Biobank dataset (see Methods) (18,19). Of the six traits investigated, total pixel-wise retinal thickness (18) and the thickness of the photoreceptor OS (19) were significantly colocalized with the RPD risk region (PPH4=0.99) (Fig. 5). The negative effect size of the RPD risk allele in both total and photoreceptor OS thickness (18,19) suggests reduced thickness associated with this SNP. Unlike the photoreceptor OS, the thickness of the other two photoreceptor cell layers IS and ONL showed little overlap with the RPD risk region (PPH4=0.58 and 0.26, respectively) (Fig. 5).

Colocalization with retinal edQTLs and the RPD risk locus showed a posterior probability of 0.92 while retinal mQTL and RPD risk locus showed the lowest overlap (posterior probability of 0.03) with the RPD risk locus (Supplementary Fig. S15) suggesting that the biological mechanism underpinning the chromosome 10 risk variant is not a methylation mediated but transcriptionally regulated, possibly via RNA editing, an alternate post-transcriptional modification to methylation. Colocalization analysis demonstrated that the RPD-risk haplotype coincided with total retinal thickness in the parafoveal region of the retina (Supplementary Fig. S16).

The consistent association of the RPD-risk lead SNP (rs11200638) across multiple retinal traits underscores its potential role in influencing retinal microstructure and in predisposing individuals to RPD, with implications for further investigations to understand disease mechanisms.

## Discussion

RPD are a clinically important AMD sub-phenotype with greater prevalence in late AMD and strong predictive value for progression to vision-threatening disease (20). Our large sample size of well-phenotyped individuals with AMD, with and without RPD, has allowed us to conduct the first robust multi-cohort RPD-specific GWAS. Our results pinpoint an AMD risk-associated region on chromosome 10 near two well-known protein-coding genes *HTRA1*, *ARMS2* and a recently annotated lncRNA gene, *BX842242.1,* all previously implicated in AMD risk (17,21), and excluded an association with the well-established chromosome 1 *CFH* AMD risk locus. Several candidate gene studies have reported the association of *ARMS2/HTRA1* locus with RPD, however, findings are inconsistent, potentially due to small cohorts, and RPD being determined before OCT imaging was available (20,22,23). Our findings appear to be further supported by a small sample GWAS from Schwarz *et al* (24) (66 RPD cases) which was underpowered to detect genetic risk for RPD in a case/control test, but did associate the same lead SNP (rs11200630) with increasing RPD load.

The inconsistent inclusion of *BX842242.1* in published retinal RNAseq and eQTL datasets may account for its more recent discovery as an AMD risk gene and now also as an RPD risk gene. We re-analysed the two largest available RNA cohorts for post-mortem retina to examine the genetic links between RPD, AMD and gene expression at this locus, as well as describing the broadly conserved genetic architecture underpinning these traits and gross retinal thickness and photoreceptor layer thickness. The complement cascade gene family on chromosome 1, comprising CFH and CFH-related proteins, has also been reported to be associated with RPD (20,22,23). However, our well-powered RPD-specific GWAS results show no association with the complement factor related genes.

The absence of association with the *CFH* gene family suggests a specific genetic pathway for the etiology of RPD that involves the *ARMS2/HTRA1* locus. The role of *ARMS2/HTRA1* and more specifically the *BX842242.1* gene in the retina is not known but a range of potential mechanisms have been suggested for *HTRA1* such as involvement in the extracellular matrix (ECM) pathway, or in promoting cellular senescence in RPE cells by impairing mitochondrial functions (25,26). However, despite extensive investigations into the *HTRA1* mechanism of action, little is known about its role in the human retina.

Multiple protein-coding and non-coding SNPs exist in the RPD risk region (Supplementary Fig. S17). For instance, SNP rs10490924 is a nonsynonymous change (A69S) located in the coding region of *ARMS2* and is shown to be likely damaging by epigenetic and mitochondrial mechanisms of actions in AMD (25,26). This SNP is in high LD with the RPD risk lead SNP (r^2^=0.98). The biological functions of coding and non-coding SNPs within the RPD risk region have been extensively studied in AMD (25,26). The RPD-risk lead SNP (rs11200638) is located within a super-enhancer/promoter region of both *HTRA1* and lncRNA *BX842242.1*, and eQTL results from this and other studies implicate induction of the lncRNA in the retina (27) (Supplementary Fig. S14). Using WGS data, we show enrichment of an insertion/deletion polymorphism (del443ins54) in participants with an extreme RPD phenotype. This structural variant is in high LD with RPD-risk lead SNP (rs11200638) as well as A69S and located in 3′UTR of ARMS2 and within the intronic region of lncRNA *BX842242.1* and involved in the destabilisation of ARMS2 mRNA (25,26). The functional role of this insertion/deletion on the lncRNA *BX842242.1* is yet to be studied.

*BX842242.1* is a recently annotated non-coding gene located antisense and upstream of *HTRA1* and may be involved in its regulation. Indeed, the in-*cis* regulatory role of lncRNAs antisense to protein-coding genes is reasonably established (28). Targeted experimentation is now required to understand the functional interaction (if any) between *HTRA1* and *BX842242.1*, especially considering their inconsistent correlation in abundance across retinal tissues and study cohorts (Supplementary Fig. S13).

Our findings combined with UK Biobank summary data from large-scale GWAS of sub-cellular retinal thickness, identified shared retinal microstructure risk loci, which pinpointed the photoreceptor outer segments of the retina as likely affected by RPD. In addition, the same data, when analyzed to examine fine-scale spatial retinal thickness association signals, uncovered the parafoveal region -a common site of RPD deposits - as having shared genetic etiology with the ARMS2/*HTRA1* locus (18). Interestingly, recent findings in an AMD cohort showed that reduced retinal thickness is often observed outside the fovea, especially the outer superior subfield of the ETDRS grid, which correlates with the areas where RPD develops over time, with the central subfield having the least involvement (29).

To address the current limitations in studying RPD (6), we used cohorts where OCT was primarily used to detect RPD (30). Although this has allowed for more accurate detection of RPD, we acknowledge that revealing genetic risk factors of RPD is not straightforward due to inconsistent definitions of RPD. Therefore, we followed a meta-analysis approach with a stringent quality control methodology that accounts for possible effects caused by variations in the RPD definition. An alternative approach would use the extent of RPD, measured in a robust, high throughput manner, likely via AI approaches applied to OCT imaging data, in individuals as a quantitative trait. Using RPD extent data, available on only a subset of our cohorts, we observed that participants in higher deciles of RPD extent are more likely to carry the RPD risk SNP. The genetic risk factors underpinning RPD quantity specifically are a target for future work including examining the rate of disease progression.

This work included European populations, however since population-specific frequencies influence disease risk it may not translate to non-European populations. Specifically, the frequency of the allele of RPD risk SNP (rs11200638) is higher in East Asians (0.4) and South Asians (0.3) than in Europeans (0.2) (based on gnomAD v3.2.1). The genetic underpinnings of RPD may be translatable across other populations with similar risk allele frequencies (Africans/Mixed Americans); however caution is needed when generalizing GWAS findings due to differing genetic factors and environmental contexts (31).

Given the nature of the RPD-specific GWAS within AMD, it is inevitable to investigate the genetic loci in concurrence with AMD. Thus, the genetics of RPD, independently of AMD, remains largely unexplored. Whether the presence of RPD is a secondary representation of the genetic risk of AMD or an independent biological mechanism(s) remains unanswered. This can be addressed by studying cohorts with isolated RPD free of retinal diseases, however such cases are relatively rare.

In summary, we present here the first evidence of specific genetic variants associated with RPD susceptibility which survive stringent genome-wide significance thresholding. We present multiple lines of evidence and a potential biological mechanism underlying genetic risk factors. Further research is needed to systematically validate our findings and to assess the functional consequences of the identified genetic variants in the retina of AMD participants more susceptible to developing RPD.

## Methods

### Ethics

In this study, prior to data collection, comprehensive ethical approvals were obtained from all relevant institutional review boards or ethics committees (Supplementary Table S15). The study was conducted in accordance with the International Conference on Harmonization Guidelines for Good Clinical Practice and the tenets of the Declaration of Helsinki, and all enrolled participants provided written informed consent. Each ethical agreement was approved by the Walter and Eliza Hall Institute of Medical Research Human Research Ethics Committee (HREC, project number 20/13LR) where the final analysis was performed.

### Study cohorts

Fourteen international cohorts joined the RPD consortium as they were able to contribute cohorts of AMD and healthy control participants 50 years of age or older, who had multimodal retinal imaging performed in both eyes to determine AMD and RPD status as well as DNA for genotyping. A list of cohorts involved in the RPD consortium is described in Supplementary Table S1. Individual sites used a combination of multimodal retinal imaging including optical coherence tomography (OCT), fundus autofluorescence (FAF), near infra-red (NIR) and color fundus photography (CFP), which were performed based on site-specific protocols (Supplementary Table S1). Retinal image grading was performed by each site to determine AMD and RPD status, whose criteria are described in Supplementary Table S1. Individual-level data was available for ten cohorts, and summary statistics were available for a further four (Supplementary Table S2).

### Harmonization of AMD and RPD phenotyping

Phenotyping across the 14 cohorts was harmonized by following rigorous person-based AMD and RPD phenotyping protocols. For individual-level data sets, this was performed at CERA and allowed a final categorization of participants into a status of AMD+ or AMD-and RPD+ or RPD-(or were excluded from the analysis if unable to categorize) across the cohorts. For data sets providing summary statistics, categorization was done by site-based graders based on the analysis plan provided by the CERA.

The AMD status (presence or absence) and stage for each participant was determined according to the Beckman Classification criteria (32). Healthy controls were required to have either no signs of aging or normal signs of aging in both eyes, while AMD stage was classified on a per person basis into i) early or intermediate AMD ii) late-stage geographic atrophy (GA) or iii) late stage neovascular AMD (nAMD) (32).

The RPD status (presence or absence) was defined in all cohorts based on OCT findings, except for one cohort (Genentech) where FAF/NIR imaging was primarily used with OCT used only for cases of questionable RPD (Supplementary Table S1). The definition of RPD+ on OCT varied across cohorts in the number of definite RPD needing to be present and varied with regard to whether confirmation on *en face* images (FAF, NIR or CFP) was required (Supplementary Table S1).

For the WGS analysis, 44 cases of AMD+/RPD++ individuals from the CERA cohort were selected with ‘extreme phenotype’ grading where the ratio of RPD to conventional drusen was >90%.

### Genotype data and quality control

Of the 14 international cohorts, DNA samples of seven cohorts extracted from blood were genotyped on the Infinium Global Screening Array-24 (GSA array v.3) (https://www.illumina.com/products/by-type/microarray-kits/infinium-global-screening.html). The remaining seven cohorts were genotyped using various SNP-chip platforms or by WGS methods within the cohort country of origin or within Europe, for the European cohorts (Supplementary Table S2 & S4). Ten cohorts provided individual level data, while four cohorts shared the GWAS summary statistics with the RPD consortium. The summary statistics cohorts were analyzed using the same shared, or as close as possible, genotyping/phenotyping quality control and analysis pipeline (Supplementary Table S1 & S2). Details of genotyping platforms in each cohort are described in Supplementary Table S4.

To ensure consistent quality control measures, an analysis plan was followed uniformly across each cohort. Detailed guidelines outlining the analytical plan for this study were shared with analysts across all four cohorts and the summary statistics were shared with the RPD Consortium. This included the removal of low-quality SNPs or samples with high missing rates of > 98% (this threshold was> 95% in the NICOLA cohort), and those SNPs exhibiting deviation from HWE (*P-values* < 5e-7 in all cohorts except for NICOLA and Genentech cohorts with P-values < 5e-6). In addition, we excluded ambiguous SNPs, non-biallelic SNPs, SNPs associated with batch effect and SNPs with MAF of < 1%. We transferred all variant identifiers to unique variant names consisting of chromosome, base position (based on hg37) and allele codes in reference and alternative alleles, respectively. Quality control procedures were implemented to ensure data accuracy for each cohort and are summarized in Supplementary Table S2.

King software version 2.3.0 (released on October 10, 2022) or PLINK v.1.0/v.2.0 (33) were used to calculate PCs and relatedness (34). Related individuals were removed in four cohorts, whereas the remaining ten cohorts included related individuals. A linear mixed model was used to perform the association analysis to deal with the related samples in ten cohorts where related individuals were included in the GWAS (Supplementary Table S2). To determine ancestries compared to the 1000 Genome reference panel, King software version 2.3.0 (released on October 10, 2022) or PLINK v.1.0/v.2.0 (33) were used. Duplicated samples, which could also be monozygotic twins, and non-European individuals were removed (Supplementary Fig. S18a).

We then imputed genotypes of nine cohorts with individual-level data to the Haplotype Reference Consortium (HRC) reference panel using the Minimac 3.0 or 4.0 pipeline via the Michigan Imputation Server or the TopMed Server. Variants were filtered to include those with high imputation quality (INFO scores >=0.7). Details of imputation reference and analysis method in each cohort are described in Supplementary Table S4.

### GWAS and meta-analysis

Given differences in phenotyping and genotyping methods and to avoid any cohort-based effects, GWAS was performed individually on each cohort. Standard or linear mixed models, using Regenie tool v3.0 (35) and the RVTESTS (36) tools, respectively, were used for the GWAS analysis (Supplementary Table S16). Age (at the date of the latest imaging) and sex were incorporated as covariates to adjust for the demographic characteristics of participants. Two to five genetic PCs, depending on cohort-specific properties, were included in the association analyses to adjust for population stratification (Supplementary Fig. S18b & Supplementary Table S16). This approach was adopted to mitigate the potential impact of phenotyping variability on the outcomes of GWAS. Details of the model used in each cohort are described in Supplementary Table S16.

A meta-analysis was performed on the GWAS results of all fourteen cohorts using METAL software (11). Additional analyses were performed to identify signals on the sex chromosome X for the eight cohorts genotyped on the GSA array v.3 (Supplementary Table S4). The genotypes of chromosome X were imputed using the HRC. The pseudo-autosomal region (PAR) was imputed jointly for males and females with the non-pseudo-autosomal region (non-PAR) being imputed separately for males and females. After separate imputation of the PAR and non-PAR regions, the association analysis was performed using the RVTESTS tool.

GWAS summary statistics for all data sources were processed through a standardized quality control pipeline (37). To ensure the robustness and reliability of our findings, we conducted additional QC on the meta-analysis results. We only retained variants for further analysis that met stringent criteria: (i) variants with a total effective sample size (Neff) exceeding >70% of the total populations, and (ii) annotated in HRC reference panel (hg19) for chromosome and position. We excluded variants with low minor allele count (MAC < 10, calculated as MAC = 2*Neff*MAF, with Neff being the effective sample size, Neff=Rsq *2/((1/NCases)+(1/NControl)) and MAF being the minor allele frequency, low imputation quality (Rsq < 0.7) or large standard error of the estimated genetic effect (SE > 10).

Further, we used RVTESTS to perform non-additive association models including recessive and dominant associations of genetic variants with RPD. The latter was performed to explore non-linear effects that might be associated with RPD risk.

We used the Genetic Association Study Power Calculator web tool (38) to compute the power of our RPD-GWAS in detecting chromosome 1 signals.

### Conditional GWAS

To investigate whether the significant region identified in the RPD GWAS represents more than one haplotype, we conducted conditional analyses using the Regenie tool v3.0 (35). Conditional GWAS was performed in cohorts genotyped on a GSA array v.3 (Supplementary Table S4).

### Post-GWAS analyses

Post-GWAS analyses, including WGS, eQTL, and PheWAS investigations, were conducted to further elucidate the genetic underpinnings of RPD phenotype associations identified in the study.

### Visual examination of associated regions

We used the LocusZoom software (39) to visualize and annotate the genomic region associated with RPD including SNPs that are in LD with top RPD risk variants derived from our summary statistics GWAS. Genes were annotated using the hg37 genome build. lncRNA *BX842242.1* was added manually.

### RPD lesion extent measurements

The extent of RPD present in an eye was determined by first segmenting individual RPD lesions within an OCT B-scan, a single image slice within a three-dimensional OCT volume scan. This was performed using an instance-based deep learning segmentation model, and it was used to derive the one-dimensional label of RPD presence along each A-scan (vertical column of pixels within a B-scan). The percentage of the A-scans within the OCT volume scan with this RPD label was thus derived to provide a two-dimensional, en face extent of RPD (12).

### WGS

Following the GWAS, a subset of individuals with extreme RPD status was selected for WGS. This subset was chosen to maximize the chances of discovering rare variants causing RPD. Additionally, the WGS data was used to explore the haplotype(s) within the RPD risk region. WGS was performed using the NovaSeq PE150 (PCR-free library preparation) aiming for 30x coverage. The raw sequence data of 44 samples (sequenced in two batches) with extreme RPD was provided as unmapped FASTQ files which were mapped to the human reference genome (based on hg37 genome assembly version) using an in-house pipeline based on GATK best-practice guidelines (40). The joint-called variants were output to variant caller format (VCF) files for: i) rare variants, small insertions and deletions (indels) in genes within the *ARMS2/HTRA1* locus, and ii) structural variants located in the chromosome 10 risk region.

Variant calling, quality control, and prioritization were carried out using an in-house pipeline by excluding variants with MAF < 0.001, including SNPs with Variant Effect Predictor (VEP) impact of moderate or higher, excluding variants predicted/annotated as benign by SIFT (41), Polyphen (42) and ClinVar tools (43). The structural variants (SV) quality control was restricted to SVs with MAF > 0.001 in the *ARMS2/HTRA1* gene region using two SV detection tools including Smoove version 0.2.8 (44) and Manta (45).

Next, an association between the RPD phenotype and the RPD risk lead SNP was evaluated using logistic regression models in AMD-/RPD-controls compared to three cohorts including: 1) Extreme RPD cases (AMD+/RPD++), 2) AMD cases with RPD (AMD+/RPD+) and 3) AMD cases with no RPD (AMD+/RPD-). These models were adjusted for age, sex, and two genetic PCs to account for population stratification, ensuring that the observed associations were not confounded by demographic or population structure factors.

### RPD risk haplotype characterization

To identify risk haplotypes associated with RPD, we conducted a haplotype analysis using high-resolution WGS genotyping data. We selected SNPs within the candidate region (Chr 10:124,205,544-124,235,544, based on hg19) that encompass a window of 15 kb of the lead variant associated with RPD. Individuals homozygous for RPD risk lead variant were chosen (N=15) for this analysis. Haplotype construction was performed firstly by phasing the genotype using PLINK v.1.9 (33). We included both common and rare variants (MAF > 0.001) to capture the full spectrum of genetic variation. Haplotype blocks were defined for variants with no missing data based on LD patterns (pairs of variants within 200 kb of each other are considered), and no frequency cut-off was set in the analysis.

### eQTL analysis

Post-mortem donor retina RNA sequencing libraries with matched genotype data were available for 404 samples (310 AMD + 94 controls) generously provided by Prof Anand Swaroop hereafter ‘NEI eQTL cohort’) (16); and up to four tissues (neural retina and RPE, peripheral and macula regions) from at least one eye from an independent cohort of 119 donors (22 AMD and 97 controls), generously provided by Dr Luz Orozco (‘Genentech cohort’) (15). Raw reads were mapped to the human reference genome GRCh38.p14 (v110; Apr 2022) using STAR in two-pass mode (46), and gene counts were quantified via subread featureCounts (47). Data were further quality controlled using edgeR (48) to remove lowly expressed genes (filterByExpr), normalize library sizes (trimmed-mean of M values), and compute log-counts-per-million (logCPM) values for downstream analysis. Targeted eQTL analysis was performed for each gene and tissue in the ARMS2 locus by extracting donor genotypes at the lead SNP and regressing log CPM values, correcting for available metadata as follows:

1. NEI eQTL cohort: log(CPM) ∼ risk allele dosage + age + sex + AMD Minnesota Grading System level + RNA integrity number + post-mortem interval (hours)
2. Genentech eQTL cohort: log(CPM) ∼ risk allele dosage + sex + AMD status + (1|donor ID)

Model 1) used the base R linear model and model 2) used lmerTest to incorporate a random term for donor ID given the availability of binocular data for 17% of the cohort. The tidyverse suite was used for data cleaning and manipulation, and the broom was used to generate summary statistics. Results for 1) were thresholded at p < 0.05; results for 2) were adjusted for multiple testing using the Benjamini-Hochberg Procedure and thresholded at adjusted p < 0.05. R packages ggplot2, cowplot and patchwork were used to generate QTL boxplots.

### Cross-trait analysis of RPD risk region

Phenotype association analyses were conducted to investigate the AMD+/RPD+ GWAS result relationship with retinal-related diseases, traits and clinical outcomes that have previously identified the *ARMS2/HTRA1* genes. We then explored the overlap of the RPD risk region in our RPD-specific GWAS and the GWAS summary statistics data of related traits. To obtain the summary statistics, we searched the GWAS Catalog (49) and PubMed for recent GWAS performed on retinal-related traits which were not available in the GWAS Catalog (18). Seven retina-relevant traits including total retinal thickness, the thickness of photoreceptor cell inner segment (IS), photoreceptor cell outer nuclear layer (ONL), photoreceptor cell outer segment (OS), retinal expression quantitative trait loci (eQTL), retinal editing quantitative trait loci (edQTLs) and retinal methylation quantitative trait loci (mQTL) were identified (Fig. 5 & Supplementary Fig. S16). The two studies on total retinal thickness and the thickness of photoreceptor cell segments used the UK BioBank dataset measures extracted from the OCT images. The total retinal thickness GWAS was conducted by Jackson et al. which is a high-throughput GWAS on fine-scale retinal thickness measurements by Artificial Intelligence (AI)-based methods across >29,000 points in the macula and was available in-house (18). The study on the thickness of photoreceptor cell segments used Topcon advanced boundary software to measure the layers (19).

### Colocalization

We conducted colocalization analyses using coloc version 5.2.2 (50) to assess shared genetic etiology across related traits.

### Statistical tests

We used a rank-based linear regression model to assess the relationship between the RPD extent and the genotype dosage of the RPD-risk lead SNP (rs11200638) while adjusting for age and sex as covariates. The Kruskal-Wallis test was conducted to examine the differences in the RPD extent across the genotype groups.

Statistical significance was defined as adjusted for multiple testing using the False Discovery Rate (FDR) or Benjamini-Hochberg P-value < 0.05, throughout this study unless it’s mentioned otherwise.

### Ethics and inclusion statement

This research included local researchers from 14 cohorts internationally. Local researchers from each cohort have fulfilled the criteria for authorship required by Nature Portfolio journals and have been included as authors since their participation was essential for the study design and implementation. Roles and responsibilities were agreed among collaborators ahead of the research with collaboration agreements in place covering study design, study implementation, data ownership, intellectual property and authorship. This manuscript includes findings that are locally relevant and were determined in collaboration with the local researchers. The research was not severely restricted or prohibited in the setting of the researchers, and has been approved by each local ethics committee or institutional review board (Supplementary Table S15). The research did not result in stigmatization, incrimination, discrimination or personal risk to participants and did not involve health, safety, security or other risk to researchers. Local researchers are able to continue using the data generated from their cohort for their own local research. Local research relevant to the study was taken into account in citations.

## Data Availability

All data produced in the present study are available upon reasonable request to the authors

## Acknowledgements

This work was supported by a National Health and Medical Research Council (NHMRC) of Australia Synergy grant to MB, BREA, RHG, ZW, ELF, and AP (GNT1181010). BREA is supported by an NHMRC Early Career Fellowship (GNT1157776), MB, RHG and ZW are supported by NHMRC Investigator Grants (GNT1195236, GNT1194667, GNT2008382), and AP by NHMRC Senior Research Fellowship (1154389). AS is supported by NEI Intramural Research Program (Z01EY000546). This work was also made possible through the Victorian State Government Operational Infrastructure Support and the NHMRC Independent Research Institute Infrastructure Support Scheme (IRIISS).

The Alienor cohort was funded by Laboratoires Théa (Clermont-Ferrand, France), Université de Bordeaux (Bordeaux, France), Fondation Voir et Entendre (Paris, France), Agence Nationale de la Recherche (2010-PRSP-011), CFSR Recherche (Club Francophone des Spécialistes de la Rétine), the French Ministry of Health (PHRC, 2012, PHRC12_157 ECLAIR) and CNSA (Caisse Nationale pour la Solidarité et l’Autonomie) (Paris, France). The Alienor study is based on the Three-City Study, which is conducted under a partnership agreement between the Institut National de la Santé et de la Recherche Médicale (INSERM), the Institut de Santé Publique, Epidémiologie et Développement of the University of Bordeaux and Sanofi-Aventis. The Fondation pour la Recherche Médicale funded the preparation and initiation of the study. The Three-City Study is also supported by the Caisse Nationale Maladie des Travailleurs Salariés,

Direction Générale de la Santé, Mutuelle Générale de l’Education Nationale, Institut de la Longévité, Regional Governments of Aquitaine and Bourgogne, Fondation de France, Ministry of Research-INSERM Programme “Cohortes et collections de données biologiques”, French National Research Agency COGINUT ANR-06-PNRA-005 and ANR 2007LVIE 003, the Fondation Plan Alzheimer (FCS 2009–2012), the Caisse Nationale pour la Solidarité et l’Autonomie (CNSA), and Roche Pharma. AM is supported by the French National Research Agency (ANR) grants ANR-23-CE12-0029-01, ANR-22-NEU2-0004-02 and Fondation Vaincre Alzheimer generic grant. Computations were performed on the Bordeaux Bioinformatics Center (CBiB) and the CREDIM computer resources, University of Bordeaux (funding provided to Pr. Stéphanie Debette by the Fondation Claude Pompidou).

The AREDS2_OCT and NEI_research studies were supported by the intramural program funds and contracts from the National Eye Institute/National Institutes of Health (NEI/NIH), Department of Health and Human Services, Bethesda, MD. Contract No. HHS-N-260-2005-00007-C. ADB Contract No. N01-EY-5-0007. Funds were generously contributed to these contracts by the following NIH institutes: Office of Dietary Supplements (ODS), National Center for Complementary and Alternative Medicine (NCCAM) now known as National Center for Complementary and Integrative Health (NCCIH), National Institute on Aging (NIA), National Heart, Lung and Blood Institute (NHLBI), and National Institute of Neurological Disorders and Stroke (NINDS).

The DUKE FEATURE study was supported by the National Eye Institute, National Institutes of Health, Bethesda, Maryland (grant no.: K23EY026988 [to E.M.L.]); F. Hoffmann La Roche Ltd., Basel, Switzerland (E.M.L. through Duke University).

The Genentech/Roche GWAS cohort (CHROMA and SPECTRI studies) and whole genome sequencing cohort were funded by F.Hoffman-LaRoche Ltd.

The Hadassah study was supported by an Israel Science Foundation grant (#3485/19).

The MACUSTAR project has received funding from the Innovative Medicines Initiative 2 Joint Undertaking under grant agreement No 116076. This Joint Undertaking has received support from the European Union’s Horizon 2020 research and innovation program and EFPIA. This communication reflects the author’s view and neither IMI nor the European Union nor EFPIA are responsible for any use that may be made of the information contained therein.

The NICOLA study received grant support from Bayer and Novartis for retinal grading.

## Additional Contributions

We are grateful to all the research participants, and the whole Reticular Pseudodrusen Consortium (Supplementary information), which includes co-ordinators, medical staff, nursing staff, research scientists, clerical staff, computer and laboratory technicians, managers and receptionists.

## Authorship Statement

All authors contributed to the work including data interpretation, drafting and approving the manuscript submission. RHG, MB, BREA, ZW, AP, ELF conceived and designed the study; CJA and RHG recruited the cohorts; CJA, ZW, RHG, EML, EAK, SFauser, UFOL, AStockwell, BLY, IC, LT, FKC, SM, DMS, KB, AT, EC, CC, RH, AM, CH, AIdH, IM, PHG, LAltay, VS, RPF, JT, MMM, FGH, CD, MND, AM, LArnould, MACUSTAR consortium, NICOLA consortium provided the GWAS datasets; RHG, ZW, CJA prepared the phenotyping plan; ZW performed the phenotyping coding; ZW, RHG, HK, AL, YB, SS provided the RPD extent algorithm; ZW, RHG, HK, CJA, LABH, FKC, DMS provided the RPD extent dataset; SFarashi, BREA, MB prepared the genetic analysis plan; SFarashi, BREA, MB, AStockwell, IM, AM, performed genetic analyses; SFarashi performed the final meta-analysis; SFarashi, JH, LWS performed the WGS analysis pipeline; LDO, SC, ASwaroop provided bulk RNAseq data and matched genotyping; BREA re-processed bulk RNAseq and ran targeted eQTL analysis; MB oversaw the genetic analysis; SFarashi, MB, BREA, RHG, ZW, AP, ELF, CJA provided detailed data interpretation and wrote the first draft.

