## Supplementary Tables for "Genetic Risk of Reticular Pseudodrusen in Age-Related Macular Degeneration: *HTRA1*/lncRNA *BX842242.1* dominates, with no evidence for Complement Cascade involvement"

### Supplementary Data

**Reticular Pseudodrusen Consortium members**

**Principal Investigators:** Robyn H Guymer, Alice Pébay, Erica L Fletcher, Melanie Bahlo, Brendan RE Ansell and Zhichao Wu

**Project manager:** Carla J Abbott

**Genetic cohorts:** Elvira Agron, Lebriz Altay, Ella Arnon, Louis Arnould, Mary Attia, Bjorn Bakker, Konstantinos Balaskas, Melinda Cain, Emily Caruso, Usha Chakravarthy, Jason Charng, Attiqa Chaudhary, Fred K Chen, Shang-Chih Chen, Emily Chew, Sae Cho, Itay Chowers, Linda Clarke, Audrey Cougnard-Grégoire, Catherine Creuzot Garcher, Catherine Cukras, Stéphanie Debette, Cécile Delcourt, Marie-Noëlle Delyfer, Anneke I den Hollander, Layal El Wazan, Sarah Elbaz-Hayun, Sascha Fauser, Daniela Ferrara, Robert P Finger, Pierre-Henry Gabrielle, Javier Gayan, Erin Gee, Emily Glover, Kai Lyn Goh, Michelle Grunin, Rachael Heath Jeffery, Catherine Helmer, Wilson Heriot, Claire Hill, Lauren AB Hodgson, Ruth Hogg, Frank G Holz, Carel Hoyng, Amy Kalantary, Haya Kashtan, Pearse A Keane, Frank Kee, Tiarnan Keenan, Samer Khateb, Meme Ko, Jean-François Korobelnik, Adi Kremer, Himeesh Kumar, Eleonora M Lad, Mélanie Le Goff, Yara Lechanteur, Jackson Lee, Rivkah Lender, Jaime Levi, Sandra Liakopoulos, Timing Liu, Ulrich FO Luhmann, Chi D Luu, Matthias M Mauschitz, Amy J McKnight, Samuel McLenachan, Aniket Mishra, Ismail Moghul, Tunde Peto, Nikolas Pontikos, Anna Rautanen, Batya Rinsky, Danial Roshandel, Danuta M Sampson, Sean Santiago, Tina Schick, Cédric Schweitzer, Merav Shiryon, Michal Shpigel, Yahel Shwartz, Vasilena Sitnilska, Laura Smyth, Amy Stockwell, Jan H Terheyden, Nikita Thomas, Liran Tiosano, Adnan Tufail, Claire Weber, Brian L Yaspan, MACUSTAR Consortium, NICOLA Consortium.

**Other researchers:** Yelena Bagdasarova, Roberto Bonelli, Steven Clarke, Maciej Daniszewski, Coen De Vente, Samaneh Farashi, Una Greferath, Satya Gunnam, Jenna Hall, Jiru Han, Marco Herold, Alex Hewitt, Victoria E Jackson, Aaron Lee, Helena Liang, Grace Lidgerwood, Jessica Ma, Luz D Orozco, Joseph Powell, Matt Rutar, Clarisa Sánchez, Roy Schwartz, Liam W Scott, Manisha Shah, Kaylene Simpson, Scott Song, Anand Swaroop, Kirstan Vessey.

**MACUSTAR consortium members:** H. Agostini, I. D. Aires, L. Altay, R. Atia, F. Bandello, P. G. Basile, J. Batuca, C. Behning, M. Belmouhand, M. Berger, A. Binns, C. J. F. Boon, M. Böttger, J. E. Brazier, C. Carapezzi, J. Carlton, A. Carneiro, A. Charil, R. Coimbra, D. Cosette, M. Cozzi, D. P. Crabb, J. Cunha-Vaz, C. Dahlke, H. Dunbar, R. P. Finger, E. Fletcher, M. Gutfleisch, F. Hartgers, B. Higgins, J. Hildebrandt, E. Höck, R. Hogg, F. G. Holz, C. B. Hoyng, A. Kilani, J. Krätzschmar, L. Kühlewein, M. Larsen, S. Leal, Y. T. E. Lechanteur, D.

Lu, U. F. O. Luhmann, A. Lüning, N. Manivannan, I. Marques, C. Martinho, A. Miliu, K. P. Moll, Z. Mulyukov, M. Paques, B. Parodi, M. Parravano, S. Penas, T. Peters, T. Peto, S. Priglinger, R. Ramamirtham, R. Ribeiro, D. Rowen, G. S. Rubin, J. Sahel, C. Sánchez, O. Sander, M. Saßmannshausen, M. Schmid, S. Schmitz-Valckenberg, J. Siedlecki, R. Silva, E. Souied, G. Staurenghi, J. Tavares, D. J. Taylor, J. H. Terheyden, A. Tufail, P. Valmaggia, M. Varano, A. Wolf, N. Zakaria

**NICOLA consortium members:** R. Hogg, A. R. Hunter, F. Kee, B. McGuinness, J. McKnight, A. Scott, J. Woodside, I. Young.

**Group-specific Acknowledgements:**

We are grateful to all the participants of the NICOLA Study, and the whole NICOLA team, which includes nursing staff, research scientists, clerical staff, computer and laboratory technicians, managers and receptionists.

We thank NIHR BioResource volunteers for their participation, and gratefully acknowledge NIHR BioResource centres, NHS Trusts and staff for their contribution. We thank the National Institute for Health and Care Research, NHS Blood and Transplant, and Health Data Research UK as part of the Digital Innovation Hub Programme. The views expressed are those of the author(s) and not necessarily those of the NHS, the NIHR or the Department of Health and Social Care.

### Supplementary Tables

**Supplementary Table S1.** RPD phenotyping protocol for each cohort.

| Cohort | RPD definition | | | | |
|---|---|---|---|---|---|
| | Present (+) | Absent (-) | Unknown | Questionable | Device |
| ALIENOR | ≥5 RPD lesions on OCT ± confirmed on en face imaging (1) | 0-4 RPD lesions on OCT | Excluded | Excluded | Heidelberg Spectralis |
| AREDS2_OCT | ≥5 RPD lesions on OCT | 0-4 RPD lesions on OCT | Excluded | Excluded | Bioptigen |
| CERA | ≥5 RPD lesions on OCT confirmed on en face imaging (1) | 0-4 RPD lesions on OCT | Excluded | Excluded | Heidelberg Spectralis |
| Duke_FEATURE | ≥5 RPD lesions on OCT (1) | 0-4 RPD lesions on OCT | Excluded | Excluded | Heidelberg Spectralis |
| EUGENDA | ≥5 RPD lesions on OCT confirmed on NIR imaging (1) (5) | 0-4 RPD lesions on OCT | Excluded | Excluded | Heidelberg Spectralis |
| Genentech | En face with FAF / IR (≥1 RPD lesions on OCT only for RPD Questionable) | 0 RPD lesions on OCT | Excluded | Excluded | FAF / IR (Cirrus OCT for RPD Questionable) |
| Hadassah | ≥5 RPD lesions on OCT (7) | 0 RPD lesions on OCT | In with RPD- | In with RPD- | Heidelberg Spectralis |
| LEI | ≥10 RPD lesions on OCT confirmed on NIR imaging (1) | 0-9 RPD lesions on OCT | Excluded | Excluded | Heidelberg Spectralis |
| MACUSTAR | ≥5 RPD lesions on OCT ± confirmed on NIR imaging (1) | 0-4 RPD lesions on OCT | In with RPD- | Excluded | Heidelberg Spectralis |
| Montrachet | ≥1 RPD lesions on OCT confirmed on NIR imaging (11) | 0 RPD lesions on OCT | Excluded | Excluded | Heidelberg Spectralis |
| NEI_Research | ≥1 RPD lesions on OCT confirmed by en face imaging (12) | 0 RPD lesions on OCT | Excluded | Excluded | Heidelberg Spectralis |
| NIHR_Bioresource | ≥5 RPD lesions on OCT (1) | 0-4 RPD lesions on OCT | Excluded | Excluded | Topcon 3D 2000 |
| NICOLA | ≥1 RPD lesions on OCT (14) | 0 RPD lesions on OCT | Excluded | Excluded | Heidelberg Spectralis |
| UBonn | ≥5 RPD lesions on OCT ± confirmed on NIR imaging (1) | 0-4 RPD lesions on OCT | In with RPD- | Excluded | Heidelberg Spectralis |

**Supplementary Table S2.** Genotyping data, and quality control of SNPs prior to imputation for each cohort.

| Cohort | Genotype data type available | Sample exclusion missing Rate cut-off | Related samples exclusion | Variant exclusion missing rate cut-off | Hardy-Weinberg equilibrium (HWE) outliers: cut-off | Sample heterozygosity outliers: cut-off | Tools used to calculate PCs/ relatedness |
|---|---|---|---|---|---|---|---|
| ALIENOR | Summary Statistics | > 95% | excluded (Pi_hat>0.1875) | > 98% | p<10-6 | mean.het±3*sd | EIGENSTRAT/PLINK |
| AREDS2_OCT | Individualized level data | > 98% | included | > 98% | p<5e-7 | mean.het±4*sd | PLINK/KING |
| CERA | Individualized level data | > 98% | included | > 98% | p<5e-7 | mean.het±4*sd | PLINK/KING |
| Duke_FEATURE | Individualized level data | > 98% | included | > 98% | p<5e-7 | mean.het±4*sd | PLINK/KING |
| EUGENDA | Individualized level data | > 98% | included | > 98% | p<5e-7 | mean.het±4*sd | PLINK/KING |
| Genentech | Summary Statistics | > 98% | excluded (Z0 cutoff 0.4) | > 98% | p<10-6 | mean.het±6*sd | Admixture /PLINK |
| Hadassah | Individualized level data | > 98% | included | > 98% | p<5e-7 | mean.het±4*sd | PLINK/KING |
| LEI | Individualized level data | > 98% | included | > 98% | p<5e-7 | mean.het±4*sd | PLINK/KING |
| MACUSTAR | Individualized level data | > 98% | included | > 98% | p<5e-7 | mean.het±4*sd | PLINK/KING |
| Montrachet | Summary Statistics | > 95% | excluded (Pi_hat>0.1875) | > 98% | p<10-6 | mean.het±3*sd | EIGENSTRAT/PLINK |
| NEI_Research | Individualized level data | > 98% | included | > 98% | p<5e-7 | mean.het±4*sd | PLINK/KING |
| NICOLA | Individualized level data* | > 95% | excluded (Pi_hat>0.1875) | > 98% | p<1e-6 | > median ± 3 x IQR | PLINK/KING |
| NIHR Bioresource | Summary Statistics | > 98% | included | > 98% | p<5e-7 | no cut-off applied | PLINK, fraposa/KING |
| UBonn | Individualized level data | > 98% | included | > 98% | p<5e-7 | mean.het±4*sd | PLINK/KING |

*Imputed and QC'ed data was shared with RPD consortium through access to NICOLA local server

**Supplementary Table S3.** Overview of the demographic and clinical characteristics across study cohorts with available individual-level data.

| Cohort | Age (years, mean) | Number of Male N (%) | RPD Status Present (among AMD patients) N (%) | Total N# |
|---|---|---|---|---|
| **ALIENOR*** | 83.2 | 149 (43.4%) | 54 (36.24%) | 343 |
| **AREDS2_OCT** | 77.1 | 141 (50.7%) | 127 (45.68%) | 147 |
| **CERA** | 74.4 | 311 (34.4%) | 317 (35.07%) | 873 |
| **Duke_FEATURE** | 71.0 | 35 (34.6%) | 32 (31.68%) | 77 |
| **EUGENDA** | 72.1 | 1433 (41.2%) | 235 (6.75%) | 3361 |
| **Genentech*** | 63.9 | 2738 (49.4%) | 537 (43.24%) | 6246 |
| **Hadassah** | 80.6 | 304 (43.1%) | 396 (56.17%) | 454 |
| **LEI** | 74.4 | 126 (36.9%) | 120 (35.19%) | 297 |
| **MACUSTAR** | 71.8 | 219 (34.8%) | 149 (23.65%) | 487 |
| **Montrachet*** | 82.2 | 306 (35.8%) | 125 (32.9%) | 854 |
| **NEI_Research*** | 76.1 | 27 (41.5%) | 13 (20.0%) | 55 |
| **NICOLA** | Not available | Not available | 141 (28.7%) | 492 |
| **NIHR Bioresource*** | 73.9 | 41 (35.6%) | 62 (53.45%) | 116 |
| **UBonn** | 75 | 80 (36.4%) | 143 (65.0%) | 204 |

*Cohorts with summary statistics available

#Total numbers are after phenotyping and genotyping quality control steps.

**Supplementary Table S4:** SNP-chip array, phasing and imputation methods/reference panel for each cohort.

| Cohort | Genotyping Array | Phasing software | Imputation software | Reference panel |
|---|---|---|---|---|
| ALIENOR | Illumina Human 610-Quad BeadChip | Eagle v2 | Minimac 2.0# | HRC* |
| AREDS2_OCT | Whole Genome Sequencing | Eagle v2.4 | Minimac 4.0# | HRC* |
| CERA | Illumina Infinium® Global Screening Array 24 | Eagle v2.4 | Minimac 4.0# | HRC* |
| Duke_FEATURE | Illumina Infinium® Global Screening Array 24 | Eagle v2.4 | Minimac 4.0# | HRC* |
| EUGENDA | mixed (Illumina Infinium® Global Screening Array 24 and Infinium CoreExome-24 v1.1 | Eagle v2.4 | Minimac 4.0# | HRC* |
| Genentech | Whole Genome Sequencing (Illumina) | Eagle v2 | Internal imputation pipeline | HRC* |
| Hadassah | Illumina Infinium® Global Screening Array 24 | Eagle v2.4 | Minimac 4.0# | HRC* |
| LEI | Illumina Infinium® Global Screening Array 24 | Eagle v2.4 | Minimac 4.0# | HRC* |
| MACUSTAR | Illumina Infinium® Global Screening Array 24 | Eagle v2.4 | Minimac 4.0# | HRC* |
| Montrachet | Illumina Human 610-Quad BeadChip | Eagle v2 | Minimac 2.0# | HRC* |
| NEI_Research | Whole Genome Sequencing | Eagle v2.4 | Minimac 4.0# | HRC* |
| NICOLA | Infinium CoreExome-24 v1.1 | Eagle v2.3 | Minimac 3.0# | HRC* |
| NIHR-Bioresource | UK Biobank Axiom Array, version 2.1 | Eagle v2 | Minimac 4.0# | HRC* |
| UBonn | Illumina Infinium® Global Screening Array 24 | Eagle v2.4 | Minimac 4.0# | HRC* |

* Haplotype Reference Consortium release 1 (HRC r1.1 2016)

### Michigan imputation server

**Supplementary Table S5:** Number of SNPs included in the meta-analysis after post-imputation QC, lambda, and effective number of samples for each cohort.

| Cohort | Number of SNPs used in met analysis | Lambda | Neff |
|---|---|---|---|
| ALIENOR | 7,324,173 | 0.94 | 68.9 |
| AREDS2_OCT | 8,045,889 | 1.09 | 73.1 |
| CERA | 7,459,799 | 1.01 | 334.2 |
| Duke_FEATURE | 7,520,155 | 1.19 | 35.6 |
| EUGENDA | 7,067,373 | 1.01 | 383.4 |
| Genentech | 7,863,286 | 1.01 | 609.6 |
| Hadassah | 7,672,138 | 1.04 | 202.4 |
| LEI | 7,521,934 | 1.03 | 131.1 |
| MACUSTAR | 7,547,158 | 1.04 | 159.6 |
| Montrachet | 8,369,653 | 0.93 | 166.9 |
| NEI_Research | 7,438,926 | 1.31 | 18.8 |
| NICOLA | 7,701,609 | 1.01 | 166.7 |
| NIHR Bioresource | 7,542,723 | 1.11 | 57.7 |
| UBonn | 6,984,307 | 1.1 | 35.6 |

**Supplementary Table S6.** Overall AMD GWAS. The lead SNP risk loci at genome-wide significant cut-off (P < 5e-8).

| Genomic Locus | Lead SNP | Chr:pos | p | Beta | Risk allele (MAF) | Identified in previous AMD GWAS |
|---|---|---|---|---|---|---|
| chr 1 | rs7514261 | 1:196700914:A:G | $3.5e^{-42}$ | -0.82 | A (0.28) | (16,17) |
| chr 10 | rs11200632 | 10:124211536:A:G | $1.98e^{-33}$ | 0.66 | G (0.36) | (16,17) |
| chr 6 | rs7772063 | 6:31906334:T:A | $4.4e^{-12}$ | -0.87 | A (0.06) | (16,17) |

**Supplementary Table S7. Genome-wide and suggestive significant signals in RPD GWAS.** One genome-wide significant locus was identified as associated with RPD in the meta-analysis GWAS. Pos = genomic location, chr= chromosome.

| rsID | chr | pos | beta | Overall MAF of the risk allele | MAF case: MAF ctrl * | Risk allele | p-value |
|---|---|---|---|---|---|---|---|
| rs11200638 | 10 | 124220544 | 0.25 | 0.6 | 0.5:0.34 | A | 3.7e-15 |
| rs76361335 | 18 | 28422097 | 0.5 | 0.01 | 0.02:0.01 | A | 3.9e-07 |

***** The average of MAF across cohorts in cases and controls.

**Supplementary Table S8:** Number of samples in GWAS1 (autosomal and chr X), GWAS2, GWAS3 and non-additive GWAS for each cohort.

| Cohort | GWAS1 (chr 1-22) | | GWAS1 (chr X) | | Conditional GWAS (chr 1-22) | | GWAS2 | | GWAS3 | | Non-additive GWAS (chr 1-22) | |
|---|---|---|---|---|---|---|---|---|---|---|---|---|
| | AMD/RPD+ | AMD/RPD- | AMD/RPD+ | AMD/RPD- | AMD/RPD+ | AMD/RPD- | AMD/RPD+ | no AMD | AMD/RPD- | no AMD | AMD/RPD+ | AMD/RPD- |
| ALIENOR | 50 | 78 | NA | NA | 50 | 78 | 50 | 215 | 78 | 215 | 50 | 78 |
| AREDS2_OCT | 68 | 79 | NA | NA | 68 | 79 | NA | NA | NA | NA | 68 | 79 |
| CERA | 306 | 377 | 306 | 377 | 306 | 377 | 306 | 190 | 377 | 190 | 306 | 377 |
| Duke_FEATURE | 30 | 47 | 30 | 47 | 30 | 47 | NA | NA | NA | NA | 30 | 47 |
| EUGENDA | 223 | 1512 | 223 | 1512 | 223 | 1512 | 223 | 1626 | 1512 | 1626 | 223 | 1512 |
| Genentech | 537 | 705 | 537 | 705 | 537 | 705 | 537 | 5004 | 705 | 5004 | 537 | 705 |
| Hadassah | 259 | 174 | 259 | 174 | 259 | 174 | 259 | 21 | 174 | 21 | 259 | 174 |
| LEI | 107 | 153 | 107 | 153 | 107 | 153 | 107 | 37 | 153 | 37 | 107 | 153 |
| MACUSTAR | 109 | 332 | 109 | 332 | 109 | 332 | 109 | 46 | 332 | 46 | 109 | 332 |
| Montrachet | 125 | 255 | NA | NA | 125 | 255 | 125 | 474 | 255 | 474 | 125 | 255 |
| NEI_Research | 12 | 43 | NA | NA | 12 | 43 | NA | NA | NA | NA | 12 | 43 |
| NICOLA | 141 | 351 | NA | NA | 141 | 351 | NA | NA | NA | NA | 141 | 351 |
| NIHR Bioresource | 62 | 54 | NA | NA | 62 | 54 | NA | NA | NA | NA | 62 | 54 |
| UBonn | 136 | 21 | 136 | 21 | 136 | 21 | 136 | 47 | 21 | 47 | 136 | 21 |
| **Total** | **2165** | **4181** | **1707** | **3321** | **2165** | **4181** | **1852** | **7660** | **3607** | **7660** | **2165** | **4181** |

**Supplementary Table S9.** GWAS2 and GWAS3 results compared to previous AMD-GWAS studies.

| Genomic Locus | Tagging SNP_GWAS2 | Tagging SNP_GWAS3 | p_GWAS2 | p_GWAS3 | Identified in previous AMD GWAS studies |
|---|---|---|---|---|---|
| chr 1 | 1:196679455:A:C | 1:196660995:A:T | 7.03E-69 | 1.95E-72 | (16,17) |
| chr 10 | 10:124215315:G:T | 10:124202126:A:T | 6.11E-85 | 1.24E-42 | (16,17) |
| chr 10 | 10:30470900:A:G | NA | 4.67E-09 | Not GW-significant | (16,17) |
| chr 10 | 10:31036512:C:T | NA | 2.61E-08 | Not GW-significant | (16,17) |
| chr 19 | 19:6718387:C:G | 19:6718387:C:G | 2.64E-13 | 5.52E-11 | (16,17) |
| chr 4 | NA | 4:177124573:A:T | Not GW-significant | 3.30E-08 | - |
| chr 6 | 6:70629849:C:T | NA | 1.82E-08 | Not GW-significant | - |
| chr 7 | 7:28155583:C:T | NA | 1.75E-08 | Not GW-significant | - |
| chr 9 | 9:136534858:A:G | NA | 1.98E-09 | Not GW-significant | - |
| chr 11 | 11:10407656:G:T | NA | Not GW-significant | 2.20E-08 | - |
| chr 12 | 12:78601978:A:G | NA | 3.40E-08 | Not GW-significant | - |
| chr 15 | 15:68884204:A:G | NA | 4.55E-09 | Not GW-significant | - |
| chr 17 | 17:49383760:A:C | NA | 1.80E-08 | Not GW-significant | - |
| chr 20 | 20:45480514:C:T | NA | 4.71E-08 | Not GW-significant | (16,17) |
| chr 21 | 21:19483180:A:T | NA | 3.50E-08 | Not GW-significant | - |

***** Only genome-wide significant results are displayed.

**Supplementary Table S10.** The union of RPD risk loci at suggestive significant cut-off (P < 1e-6) in additive and non-additive (recessive and dominant) models.

| MarkerName | P (dominant model) | BETA (dominant model) | P (recessive model) | BETA (recessive model) | P (additive model) | BETA (additive model) | Gene |
|---|---|---|---|---|---|---|---|
| **10:124227624:C:T** | 1.99e-07 | -0.23 | 8.99e-07 | -0.21 | 4.85e-15 | -0.245 | HTRA1/ARMS2/ BX842242.1 |
| **10:124229203:C:T** | 3.57e-07 | -0.23 | NaN | NaN | 1.60e-14 | -0.24 | HTRA1/ARMS2/ BX842242.1 |
| **6:125474791:T:G** | Not significant | - | 3.8e-07 | -0.22 | - | Not significant | TPD52L1/ HDDC2 |
| **7:22333231:A:G** | Not significant | - | 7.8e-07 | 0.22 | - | Not significant | RAPGEF5 |
| **7:22334012:G:C** | Not significant | - | 5.9e-07 | -0.22 | - | Not significant | RAPGEF5 |

**Supplementary Table S11.** The Wald test results compare the odds ratios between the different cohorts.

| Comparison | Wald Statistic | P-value |
|---|---|---|
| AMD/RPD++ vs AMD/RPD+ | 7.451 | 0.006339 |
| AMD/RPD++ vs AMD/RPD- | 19.753 | 0.000009 |
| AMD/RPD+ vs AMD/RPD- | 19.006 | 0.000013 |

**Supplementary Table S12:** Haplotype blocks identified within RPD risk region.

| Haplotype block | CHR | BP1 | BP2 | KB | NSNPS |
|---|---|---|---|---|---|
| H0 | 10 | 124199141 | 124206780 | 7.64 | 16 |
| H1 | 10 | 124208466 | 124208520 | 0.055 | 2 |
| H2 | 10 | 124216372 | 124223818 | 7.447 | 36 |
| H3 | 10 | 124225297 | 124225685 | 0.389 | 3 |
| H4 | 10 | 124226154 | 124226983 | 0.83 | 8 |

**Supplementary Table S13.** eQTL results for RPD-risk region, Genentech data.

| Gene | tissue | estimate | std.error | statistic | p.value | fdr |
|---|---|---|---|---|---|---|
| BX842242.1 | Retina, non-Macula | 0.210685 | 0.069966 | 3.011259 | 0.003135 | 0.037625 |
| HTRA1 | Retina, Macula | -0.15167 | 0.051023 | -2.97262 | 0.003597 | 0.039569 |
| BX842242.1 | Retina, Macula | 0.230705 | 0.078471 | 2.94001 | 0.003968 | 0.039682 |
| BX842242.1 | RPE, non-Macula | 0.345614 | 0.11852 | 2.916081 | 0.00422 | 0.039682 |

**Supplementary Table S14.** eQTL results for RPD-risk region, NEI data.

| Gene | tissue | estimate | std.error | statistic | p.value |
|---|---|---|---|---|---|
| BX842242.1 | RPE | 0.319 | 0.0455 | 7.02 | 9.76E-12 |
| HTRA1 | RPE | -0.0835 | 0.0268 | -3.11 | 1.99E-03 |
| PLEKHA1 | RPE | 0.00745 | 0.0288 | 0.259 | 7.96E-01 |

**Supplementary Table S15:** Ethics committees and approval number for each clinical cohort.

| Cohort Name | Ethics Committee / Institutional Review Board | Decision (Approved/Waived) | Ethics Number | Clinical Site / Principal Investigator |
|---|---|---|---|---|
| ALIENOR | Ethical Committee of Bordeaux (Comité de Protection des Personnes Sud-Ouest et Outre-Mer III) | Approved | 2006/10 | Not applicable |
| AREDS2_OCT | National Institute of Health Institutional Review Board | Approved | 08-EI-0043 | Not applicable |
| CERA | The Royal Victorian Eye and Ear Hospital Human Research Ethics Committee | Approved | 20/1459H; 95/283H/15; 11/1031H (Project AMD, AMDRFS, iPSC) | Not applicable |
| Duke_FEATURE | Duke University Institutional Review Board | Approved | #00036248; #00107399 | Not applicable |
| EUGENDA | Ethics committees of University Hospital Cologne, Germany and Nijmegen, Netherlands | Approved | 07-030 | Not applicable |
| Genentech | Nova Scotia Health Authority Research Ethics Board, Halifax Canada | Approved | No number; Chroma | Alan Cruess |
| Genentech | Belberry Human Research Ethics Committee, Eastwood Australia | Approved | No number; Chroma | Alan Luckie; Jennifer Arnold; Paul Mitchell |
| Genentech | Quorum, Seattle USA | Approved | No number; Chroma | Alan Ruby; Ali Tabassian, Allen Thach; Brad Baker; Brandon Busbee; Brian Berger; Carmelina Gordon; Clement Chan; Dante Pieramici; Darma Ie; David Boyer; David Callanan; David Eichenbaum; Dennis Marcus; Dilsher Dhoot; George Novalis; Glenn Stoller; Gregg Kokame; Gregory A Fox; H-Logan Brooks-Jr; Howard Fine; Ivan Suner; James Combs; Jaed Nielsen; Jeffrey Heier; Jeffrey Morre; Jeffrey Zheutlin; Karl Olsen; Laurent Lalonde; Lawrence Singerman; Mark Michels; Matthew Wood; Michael Ober; Miguel Busquets; Patrick Higgins; Paul Lee; Paul Raskauskas; Philip Ferrone; Pravin Dugel; Raj Maturi; Randy Katz; Ricky Isernhagen; Robert Hampton, Robert Johnson; Robert Kwun, Robert Stoltz; Robert Wirthlin; Robert Wong; Ryan Tarantola; Sam Mansour; Sanford Chen; Seong Lee; Soraya Rofagha; Stuart Burgess; Sunil Guta; Sunil Patel; Todd Schneiderman; Veeral Sheth; Victor Gonzalez; Vrinda Hershberger |
| Genentech | Comite de Etica en Investigacion Clinica (CEIC), Buenos Aires Argentina | Approved | No number; Chroma | Alberto Zambrano; Carlos Zeolite; Federico Furno Sola; Patricio Schlottmann |
| Genentech | Medical Research Council, Ethics Committee for Clinical Pharmacology, Budapest Hungary | Approved | No number; Chroma | Andras Seres; Attila Vajas; Balazs Varsanyi |
| Genentech | Wester International Review Board, Puyallup USA | Approved | No number; Chroma | Andre Witkin; Barbara Blodi; Richard Feist |
| Genentech | Vanderbilt University Institutional Review Board, Nashville USA | Approved | No number; Chroma | Anita Agarwal |

| | | | | |
|---|---|---|---|---|
| Genentech | Comites de protection des personnes (CPP) Ile de France V, Paris France | Approved | No number; Chroma | Bahram Bodaghi; Catherine Francais; Eric Souied; Ramin Tadayoni; Salomon Yves Cohen |
| Genentech | National Research Ethics Service (NRES) Committee North East, Newcastle & North Tyneside 1, Jarrow UK | Approved | No number; Chroma | Baljean Dhillon; Christopher Brand; Sajjad Mohmood, Sanjiv Banerjee; Tim Jackson |
| Genentech | Ethics Committee of the Hospital Zilina, Zilina Slovakia | Approved | No number; Chroma | Blandina Lipkova |
| Genentech | Komisja Bioetyczna przy Okregowej Izbie Lekarskiej w Gdansku, Gdansk Poland | Approved | No number; Chroma | Bozena Romanowska-Dixon; Dorota Raczynska; Slawomir Teper |
| Genentech | De Videnskabsetiske Komiteer for Region Hovedstade, Hillrod Denmark | Approved | No number; Chroma | Caroline Laugesen |
| Genentech | Ethik-Kommision der Medizinischen Hochschule Hannover, Hannover Germany | Approved | No number; Chroma | Carsten Framme |
| Genentech | Ethik-Kommision der Arztekammer Westfalen-Lippe, Munster Germany | Approved | No number; Chroma | Daniel Pauleikhoff; Nicole Eter |
| Genentech | Comite de Etica en Investigacion de Mexico Centre for Clinical Research SA de CV, Distrito Federal Mexico, Mexico | Approved | No number; Chroma | David Lozano Rechy |
| Genentech | University of British Columbia Clinical Research Ethics Board, Vancouver Canada | Approved | No number; Chroma | David Maberley |
| Genentech | Ce Indip. Presso la fond. Ptv Policl. Tor Vergata, Roma Italy | Approved | No number; Chroma | Federico Ricci |
| Genentech | Comitato Di Bioetica Della ASL Di Sassari, Sassari Italy | Approved | No number; Chroma | Francesco Boscia |
| Genentech | Comitato Etico Milano Area B, Milano Italy | Approved | No number; Chroma | Francesco Viola |
| Genentech | Ethikkommission an der Medizinischen Fakultat der Rheinischen Friedrick-Wilhelms-Universitat Bonn, Bonn Germany | Approved | No number; Chroma | Frank Holz |
| Genentech | Medisch Ethische Toetsingscommissie Amsterdam (METC AMC), Amsterdam The Netherlands | Approved | No number; Chroma | G. Dijkman |
| Genentech | Chesapeake Research Review Institutional Review Board, Columbia USA | Approved | No number; Chroma | Ghassan Ghorayeb |
| Genentech | Comite de Bioetica de la Clinica Anglo Americana, Lima Peru | Approved | No number; Chroma | Guillermo Reategui |
| Genentech | Eticka komisia Nomonia Sveteho Michala, Bratislava Slovakia | Approved | No number; Chroma | Hedviga Mikova |
| Genentech | Comite Etico de Investigacion Clinica de la Comunidad Autonoma del Pais Vasco (CEIC-E), Vitoria Spain | Approved | No number; Chroma | Javier Araiz |
| Genentech | Comite Etico de Investigacion Clinica, Barcelona Spain | Approved | No number; Chroma | Javier Araiz; Javier Montero; Jordi Mones; Jorge Mataix; Laura Sararols; Luis Arias |

| | | | | |
|---|---|---|---|---|
| Genentech | Lahey Clinic Institutional Review Board, Burlington, USA | Approved | No number; Chroma | Jeffrey Chang |
| Genentech | Human Subjects Research Office, Miami USA | Approved | No number; Chroma | Jorge Fortun |
| Genentech | Universiteit Gent, Commissie voor Medische Ethiek, Gent Belgium | Approved | No number; Chroma | Julie De Zaeytijd |
| Genentech | Ethikkommission an der Medizinischen Fakultat Universitat zu Koln, Germany | Approved | No number; Chroma | Lebriz Altay |
| Genentech | Comite de Etica en Investigacion Clinica (CEIC) Hospital de Bellvitge, Barcelona Spain | Approved | No number; Chroma | Luis Arias |
| Genentech | Northwestern University Office for the Protection of Research Subjects, Chicago USA | Approved | No number; Chroma | Manjot Gill |
| Genentech | Eticka Komisia pri Fakultna Nemocnica Trencin, Trencin Slovakia | Approved | No number; Chroma | Marek Kacerik |
| Genentech | Comitato Etico Centrale Sezione - Bietti Istituti Fisioterapici Ospitalieri (IFO), Roma Italy | Approved | No number; Chroma | Maria Cristina Parravano |
| Genentech | Sydney Local Health District Ethics Review Committee RPAH Zone), Newtown Australia | Approved | No number; Chroma | Mark Gillies |
| Genentech | Ethik-Kommission der Universitat Gottingen, Gottingen Germany | Approved | No number; Chroma | Nicolas Feltgen |
| Genentech | Ethikkommission des AKH Linz, Linz Austria | Approved | No number; Chroma | Nicole Schnelzer |
| Genentech | Sunnybrook Science Health Centre, Toronto Canada | Approved | No number; Chroma | Peter Kertes |
| Genentech | University of Texas Southwestern, Dallas USA | Approved | No number; Chroma | Rafael Ufret-Vincenty |
| Genentech | Wake Forest University Health Sciences Institutional Review Board, Winston-Salem USA | Approved | No number; Chroma | Rajiv Shah |
| Genentech | Academisch Medisch Centrum, Amsterdam Netherlands | Approved | No number; Chroma | Reinier Schlingemann |
| Genentech | University of California San Francisco Committee on Human Research, San Francisco USA | Approved | No number; Chroma | Robert Bhisitkul |
| Genentech | New York Eye & Ear Infirmany, Icahn School of Medicine at Mount Sinai Institutional Review Board, New York USA | Approved | No number; Chroma | Ronald Gentile |
| Genentech | EK Lubeck Institutional Review Board, Lubeck Germany | Approved | No number; Chroma | Salvatore Grisanti |
| Genentech | St Michael's Hospital Institutional Review Board, Toronto Canada | Approved | No number; Chroma | Shelley Boyd |
| Genentech | Kantonale Ethikkommission Zurich, Zurich Switzerland | Approved | No number; Chroma | Stephan Michels |

| | | | | |
|---|---|---|---|---|
| Genentech | University of California Los Angeles Office of the Human Research Protection Program, Los Angeles USA | Approved | No number; Chroma | Stephan Schwartz |
| Genentech | The Cleveland Clinic Institutional Review Board, Cleveland USA | Approved | No number; Chroma | Sumit Sharma |
| Genentech | Comite de Etica en Investigacion de la Asociacion para Evitar la Ceguera en Mexico IAP, Ciudad de Mexico, Mexico | Approved | No number; Spectri | Virgilio Morales Canton |
| Genentech | Quorum, Seattle USA | Approved | No number; Spectri | Alan Gordon; Alexamder Eaton; Allen Ho; Andrew Antosyzk; Brian Connolly; Cameron Javid; Carl Baker; Charles C Wykoff; Christine R Gonzales; Daniel Miller; Dante Pieramici; David Eichenbaum; David Saperstein; David M Brown; Deeba Husain; Eric Suan; H Logan Brooks Jr; Jared Nielsen; Joel Pearlman, Jonathan Williams; Jorge Calzada William Brides Jr; K Baily Freund; Larry Helperin; Leonard Feiner; Lloyd Clark; Mark Johnson, Mark Wieland; Mimi Liu; Nancy Holekamp; Nauman Chaudhry; Nokolas London; Paul Wishaar; Peter Pavan; Prema Abaham; Raymond Sjaarda; Richard Breazeale; Richard Dreyer; Robert C Wang; Scott Foxman; Sean Adrean; Subhransu K Ray; Sunil Patel; Virgil Alfaro; William Durant; |
| Genentech | Comite de Etica en Investigacion Clinica (CEIC) de Navarra, Dapartamento de Salud Pabellon de Docencia, Pamplona Spain | Approved | No number; Spectri | Alfredo Garcia-Layana; Francisco Gomez Ulla; Jose Manuel Ortiz; Marta Figueroa; Rafael Navarro; Ramon Torres; Roberto Gellego-Pinazo Imaz |
| Genentech | Regionala Etikprovningsamndem I Stockholm Karolinska Institute, Stockholm Sweden | Approved | No number; Spectri | Anders Kvanta |
| Genentech | Ethick-Kommission der Universitat Regensburg, Germany | Approved | No number; Spectri | Andreea Gamulescu |
| Genentech | Eticka komisia UN Bratislava, Bratislava Slovakia | Approved | No number; Spectri | Andrej Cernak |
| Genentech | National Research Ethics Service (NRES) Committee South Central-Berkshire, Bristol UK | Approved | No number; Spectri | Andrew Lotery; Andrew Browning; Clare Bailey; Faruque Ghanchi; Ian Pearce; Michel Williams; Niro Narendran; Quresh Mohamed; Richard Gale; Simona Esposti |
| Genentech | Samara State Medical University, Samara Russian Federation | Approved | No number; Spectri | Andrey Zolotarev |
| Genentech | Comissao de Etica para Investigacao Clinica, Lisboa Portugal | Approved | No number; Spectri | Angela Carneiro |
| Genentech | Belberry Human Research Ethics Committee, Eastwood Australia | Approved | No number; Spectri | Anthony Kwan; Devinder Chauhan; Fred Chen; Jagjit Gilhotra |
| Genentech | Ethikkommission der Medizinischen Fakultat, Munchen Germany | Approved | No number; Spectri | Armin Wolf |
| Genentech | Comite de Etica en Investigacion Clinica (CEIC), Buenos Aires Argentina | Approved | No number; Spectri | Arturo Alezzandrini; Mauricio Martinez Cartier |

| | | | | |
|---|---|---|---|---|
| Genentech | Medical Research Council, Ethics Committee for Clinical Pharmacology, Budapest Hungary | Approved | No number; Spectri | Balazs Varsanyi; Jonos Nemeth; Peter Vamosi |
| Genentech | Hacettepe University Medical Faculty Clinical Research Ethics Committee, Ankara Turkey | Approved | No number; Spectri | Bora Eldem; Gursel Yilmaz; Jale Mentes; Nur Kir; Osman Saatci |
| Genentech | Wills Eye Hospital Institutional Review Board, Philadelphia USA | Approved | No number; Spectri | Carl Regillo |
| Genentech | Medisch Ethische Toetsingscommissie Amsterdam (METC AMC), Amsterdam The Netherlands | Approved | No number; Spectri | Carel Hoyng |
| Genentech | Comite Etica para la Investigacion Universidad de San Martin de Porres, Lima Peru | Approved | No number; Spectri | Carlos Fernandez; Silvio Lujan |
| Genentech | Comites de protection des personnes (CPP) Sud Ouest Et Outre Mer III, Bordeaux France | Approved | No number; Spectri | Catherine Creuzot Garcher; Francois Devin; Jean Francois Korobelnik;Laurent Kodjikian; Michel Weber; Saddek Mohand Said |
| Genentech | Citta della salute e della scienza di Torino, Comitatio Etico, Torino Italy | Approved | No number; Spectri | Chiara Eandi |
| Genentech | Ethikkommission Technische Universitat Munchen, Munchen Germany | Approved | No number; Spectri | Chris P Lohmann |
| Genentech | Oregon Health and Science University Institutional Review Board, Portland USA | Christina Flaxel | No number; Spectri | Christina Flaxel |
| Genentech | University of California Los Angeles Office of the Human Research Protection Program, Los Angeles USA | Approved | No number; Spectri | David Sarraf |
| Genentech | University of Nebraska Medical Center Institutional Review Board, Omaha USA | Approved | No number; Spectri | Diana Do |
| Genentech | Duke University Health System Institutional Review Board, Durham USA | Approved | No number; Spectri | Eleanora Lad |
| Genentech | SAHI Republic Clinical Ophthalmological Hospital of Ministry of Health of Tatrstan Republic, Kazan Russian Federation | Approved | No number; Spectri | Elmira Abdulaeva |
| Genentech | Komisja Bioetyczna przy Okregowej Izbie Lekarskiej, Bydgoszcz Poland | Approved | No number; Spectri | Ewa Herba; Jakub Kaluzny; Jerzy Nawrocki; Marta Misiuk-Hojlo |
| Genentech | Comitato Etico Irccs Ospedale San Raffaele, Milano Italy | Approved | No number; Spectri | Francesco Bandello |
| Genentech | Comite Etico de Investigacion Clinica de Galicia, Subdirection Xeral de Farmacia e Produtos Sanitar, Santiago de Compostela, A Coruna Spain | Approved | No number; Spectri | Francisco Gomez Ulla |
| Genentech | Wayne State University Human Investigation Committee, Detroit USA | Approved | No number; Spectri | Gary Abrams |
| Genentech | Ethikkommission der Medizinischen Fakultat, Heidelberg Germany | Approved | No number; Spectri | Gerd Auffarth |
| Genentech | Comitato Etico Area Vasta Centra, Fireze Italy | Approved | No number; Spectri | Gianni Virgili |
| Genentech | Comitato Etico Interaziendale Milano Area A, Milano Italy | Approved | No number; Spectri | Giovanni Staurenghi |

| | | | | |
|---|---|---|---|---|
| Genentech | Etik-Kommission der Albert-Ludwigs Universitat, Frieburg Germany | Approved | No number; Spectri | Hansjurgen Agostini |
| Genentech | Univresity of California San Diego Human Research Protections Program, La Jolla USA | Approved | No number; Spectri | Henry Ferreyra |
| Genentech | Comite de Etica en Investigacion Clinica (CEIC) Complejo Hospitalario de Albacete, Albacete Spain | Approved | No number; Spectri | Jose Manuel Ortiz |
| Genentech | Comite de Etica en Investigacion de Mexico Centre for Clinical Research SA de CV, Distrito Federal Mexico, Mexico | Approved | No number; Spectri | Juan Ramirez Estudillo |
| Genentech | UZ Leuven Gasthuisberg, Commissie Medische Ethiek, Leuven Belgium | Approved | No number; Spectri | Julie Jakob |
| Genentech | Ethikkommission Nordwest- und Zentralschweiz, Basel Switzerland | Approved | No number; Spectri | Katja Hatz |
| Genentech | Ethik-Kommission der Landesarztekammer Rheinland-P, Mainz Germany | Approved | No number; Spectri | Lars-Olof Hattenbach |
| Genentech | CHU Brugmann (Victor Horta), Comite d'Etique, Broxelles Belgium | Approved | No number; Spectri | Laurence Postelmans |
| Genentech | University of California Davis Institutional Review Board, Sacramento USA | Approved | No number; Spectri | Lawrence Morse |
| Genentech | Medical University of South Carolina Office of Research Integrity Institutional Review Board, Charleston USA | Approved | No number; Spectri | Lucian Del Priore |
| Genentech | Comite de Etica en Investigacion Clinica (CEIC) Hospital Universitario La Princesa, Madrid, Spain | Approved | No number; Spectri | Marta Figueroa; Ramon torres Imaz |
| Genentech | Comitato Etico Regione Liguria (Sezione 2), Gonova Italy | Approved | No number; Spectri | Massimo Nicolo |
| Genentech | Western Institutional Review Board, Puyallup USA | Approved | No number; Spectri | Matthew Ohr; Nieraj Jain; Scott Oliver |
| Genentech | De Videnskabsetiske Komiteer for Region Hovedstade, Hillrod Denmark | Approved | No number; Spectri | Michael Larsen |
| Genentech | Comite Institucional de Etica en Investigacion de la Asociacion Benefica Prisma, Lima Peru | Approved | No number; Spectri | Miguel Guzman |
| Genentech | Comitato Etico Regionale Unico (CERU), Udine Italy | Approved | No number; Spectri | Paolo Lanzetta |
| Genentech | Johns Hopkins Medine Institutional Review Board, Baltimore USA | Approved | No number; Spectri | Peter Campochiaro |
| Genentech | University of Miami Miller School of Medicine, Human Subject Research Office, Miami USA | Approved | No number; Spectri | Philip Rosenfeld |
| Genentech | Instituto de Microcirugia Ocular Comite de Etica en Investigacion Clinica (CEIC), Barcelona Spain | Approved | No number; Spectri | Rafael Navarro |
| Genentech | Instituto Mexicano de Oftalmologia, Comite de Etica en Investigacion (CEI ), Quereta Mexico | Approved | No number; Spectri | Renata Garcia Franco |
| Genentech | Scott and White Memorial Hospital Institutional Review Board, Temple USA | Approved | No number; Spectri | Robert Rosa |

| | | | | |
|---|---|---|---|---|
| Genentech | Comite Etico de Investigacion Clinica, Hospital Universitari I Politecnic LA FE, Valencia Spain | Approved | No number; Spectri | Roverto Gellego-Pinazo |
| Genentech | Human Research and Ethics Committee Royal Victorian Eye and Ear Hospital, East Melbourne Australia | Approved | No number; Spectri | Robyn Guymer |
| Genentech | Comissao de Etica para Investigacao Clinica (CEIC), Lisboa Portugul | Approved | No number; Spectri | Rufino Silva; Sara Vaz-Pereira |
| Genentech | Kantonale Ethikkommission Bern KEK, Bern Switzerland | Approved | No number; Spectri | Sebastian Wolf |
| Genentech | Weill Cornell Medical Center Institutional Review Board | Approved | No number; Spectri | Szilard Kiss |
| Genentech | Standford University Research Compliance Office, Palo Alto USA | Approved | No number; Spectri | Theodore Leng |
| Genentech | Ethik-Kommission der Medizinischen Fakultat der Eberhard-Kars-Universitat und am Univresitatsklinikum, Tugingen Germany | Approved | No number; Spectri | Ulrich Bartz-Schmidt |
| Genentech | Ethikkommission der Universitat Wien/AKH, Wien Austria | Approved | No number; Spectri | Ursula Schmidt-Erfurth |
| Genentech | FSBI Scientific Research Institute of Eye Diseases of Russia Academy of Medical Sciences, Moscow Russian Federation | Approved | No number; Spectri | Valery Erichev |
| Genentech | University of Virginia Institutional Review Board for Health Sciences Research, Charlottesville, USA | Approved | No number; Spectri | Yevgeniy Shildkrot |
| Genentech eQTL | Pharma Repository Genernance Committee at Genentech (Genentech/Roche Institutional Ethical Committee) | Approved | No number | Not applicable |
| Hadassah | Institutional review board of the Hadassah Medical Center | Approved | #382-19 | Not applicable |
| LEI | Sir Charles Gairdner Group Human Research Ethics Committee | Approved | 2013-118 (LEAD) | Not applicable |
| LEI | University of Western Australia, Human Ethics Office of Research | Approved | 2021/ET000151 (WARD) | Not applicable |
| MACUSTAR | University Hospital Bonn Ethics Committee | Approved | 384/17 | CS015 |
| MACUSTAR | Paris Ouest IV Ethics Committee | Approved | 04/18_2 | CS003 |
| MACUSTAR | AIBILI Ethics Commitee | Approved | 032/2017/AIBILI/CE | CS001 |
| MACUSTAR | Comissão de Ética para a Saúde do CHSJ | Approved | dated 12/01/2018 | CS032 |
| MACUSTAR | London Queen Square Research Ethics Committee | Approved | 18/LO/0145 | CS010 |
| MACUSTAR | Center for Sundhed Glostrup Ethics Committee | Approved | H-18000126 | CS030 |
| MACUSTAR | Comitato Etico Milano | Approved | 37910/2018 | CS034 |
| MACUSTAR | Il Comitato Etico dell´Ospedale San Raffaele | Approved | dated 25/10/2018 | CS067 |
| MACUSTAR | Radboudumc Technology Center Ethics Committee | Approved | 2017-3954 | CS017 |
| MACUSTAR | LUMC commissie medische ethiek | Approved | L18.055/SH/sh | CS106 |
| Montrachet | Kremlin Bicetre and Dijon Ethics Committee | Approved | 2009-A00448-49 | Not applicable |

| | | | | |
|---|---|---|---|---|
| NEI_Research | National Institute of Health Institutional Review Board | Approved | CR002060 | Not applicable |
| NEI_eQTL | National Institute of Health Ethics Committee | Waived (no way to identify individuals that have donated post-mortem retina samples) | Not applicable | Not applicable |
| NIHR_Bioresource | South Central - Berkshire B Research Ethics Committee | Approved | 19/SC/0337 | Research Tissue Bank – RTB-GEN |
| NICOLA | School of Medicine, Dentistry and Biomedical Sciences Ethics Committee, Queen's University Belfast | Approved | 23/12 | Not applicable |
| UBonn | University Hospital Bonn Ethics Committee | Approved | 045/18; 513/20 | Not applicable |

**Supplementary Table S16:** Models, tools and covariates used in association analysis for each cohort.

| Cohort | Association Model | Association analysis tool | Covariates |
|---|---|---|---|
| ALIENOR | Logistic ridge regression model | REGENIE | sex, age, PC1:PC2 |
| AREDS2_OCT | linear mixed model | rvtests | sex, age, PC1:PC5 |
| CERA | linear mixed model | rvtests | sex, age, PC1:PC5 |
| Duke_FEATURE | linear mixed model | rvtests | sex, age, PC1:PC5 |
| EUGENDA | linear mixed model | rvtests | sex, age, PC1:PC5 |
| Genentech | linear mixed model | rvtests | sex, age, PC1:PC5 |
| Hadassah | linear mixed model | rvtests | sex, age, PC1:PC5 |
| LEI | linear mixed model | rvtests | sex, age, PC1:PC5 |
| MACUSTAR | linear mixed model | rvtests | sex, age, PC1:PC5 |
| Montrachet | Logistic ridge regression model | REGENIE | sex, age, PC1:PC2 |
| NEI_Research | linear mixed model | rvtests | sex, age, PC1:PC5 |
| NICOLA | linear mixed model | rvtests | sex, age, PC1:PC5 |
| NIHR-Bioresource | linear mixed model | rvtests | sex, age, PC1:PC5 |
| UBonn | linear mixed model | rvtests | sex, age, PC1:PC5 |
