## Supplementary Figures for "Genetic Risk of Reticular Pseudodrusen in Age-Related Macular Degeneration: *HTRA1*/lncRNA *BX842242.1* dominates, with no evidence for Complement Cascade involvement"

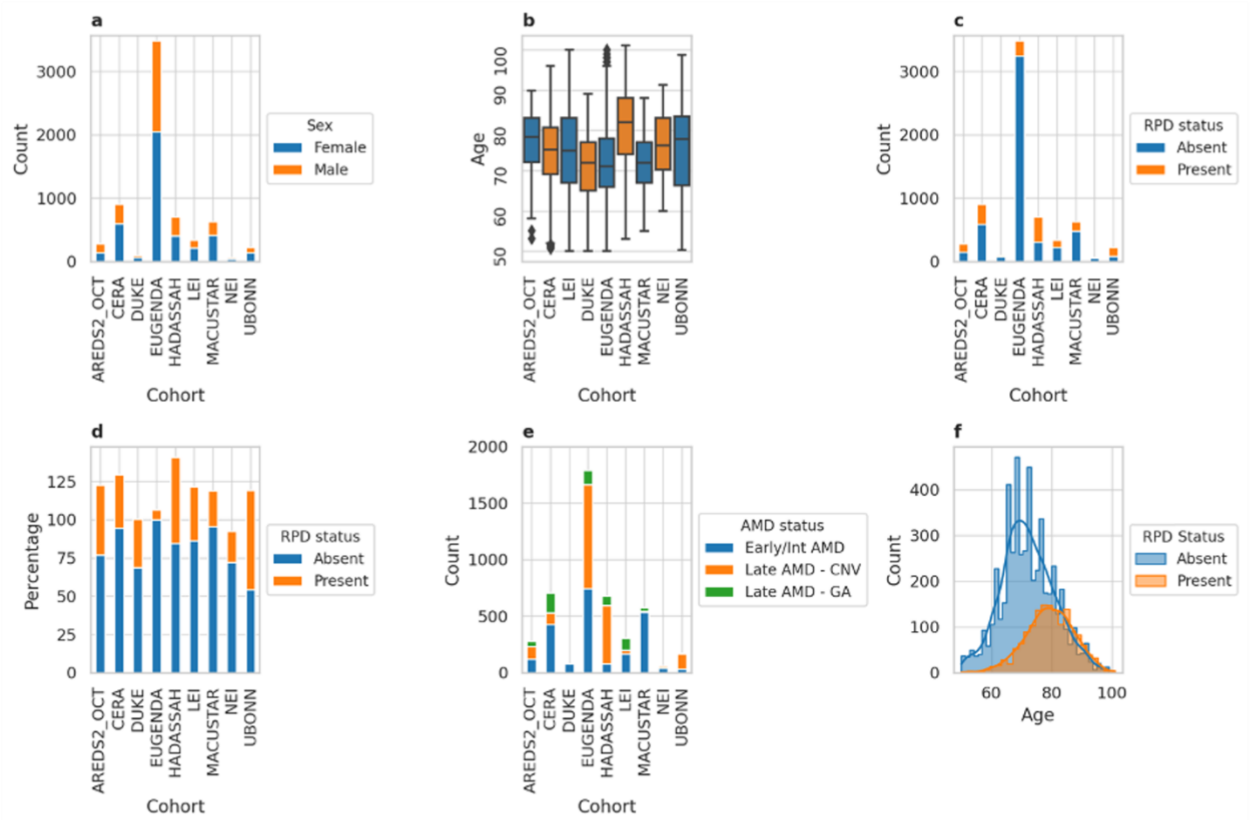


**Supplementary Figure S1. Demographic and phenotypic characteristics of the study cohorts with individual-level genotype data available**; **a** Sex distribution within different cohorts. Each cohort is represented by the number of participants within a bar plot, each denoted by a different color. The bar plots are split into male and female participants (x-axis); **b** Age distribution within different cohorts. Each cohort is represented by a different color, and density lines are overlaid to show the distribution of ages within each cohort; **c** Distribution of the RPD phenotype among study participants by cohort. The x-axis indicates RPD status, and the y-axis shows the count of participants in each category, with each cohort represented by a different color; **d** Distribution of the RPD phenotype stratified by sex. The x-axis represents the RPD status and the y-axis indicates the count of participants, with different colors representing male and female participants; **e** Distribution of the RPD phenotype stratified by AMD status. The x-axis represents the RPD status and the y-axis shows the count of participants, with different colors representing different AMD severity statuses; **f** Age distribution stratified by the RPD phenotype. The x-axis represents age, and the y-axis shows the count of participants, with different colors indicating the RPD status. Note that the demographic and phenotypic characteristics data for the NICOLA cohort is not shown in this figure due to the unavailability of data.

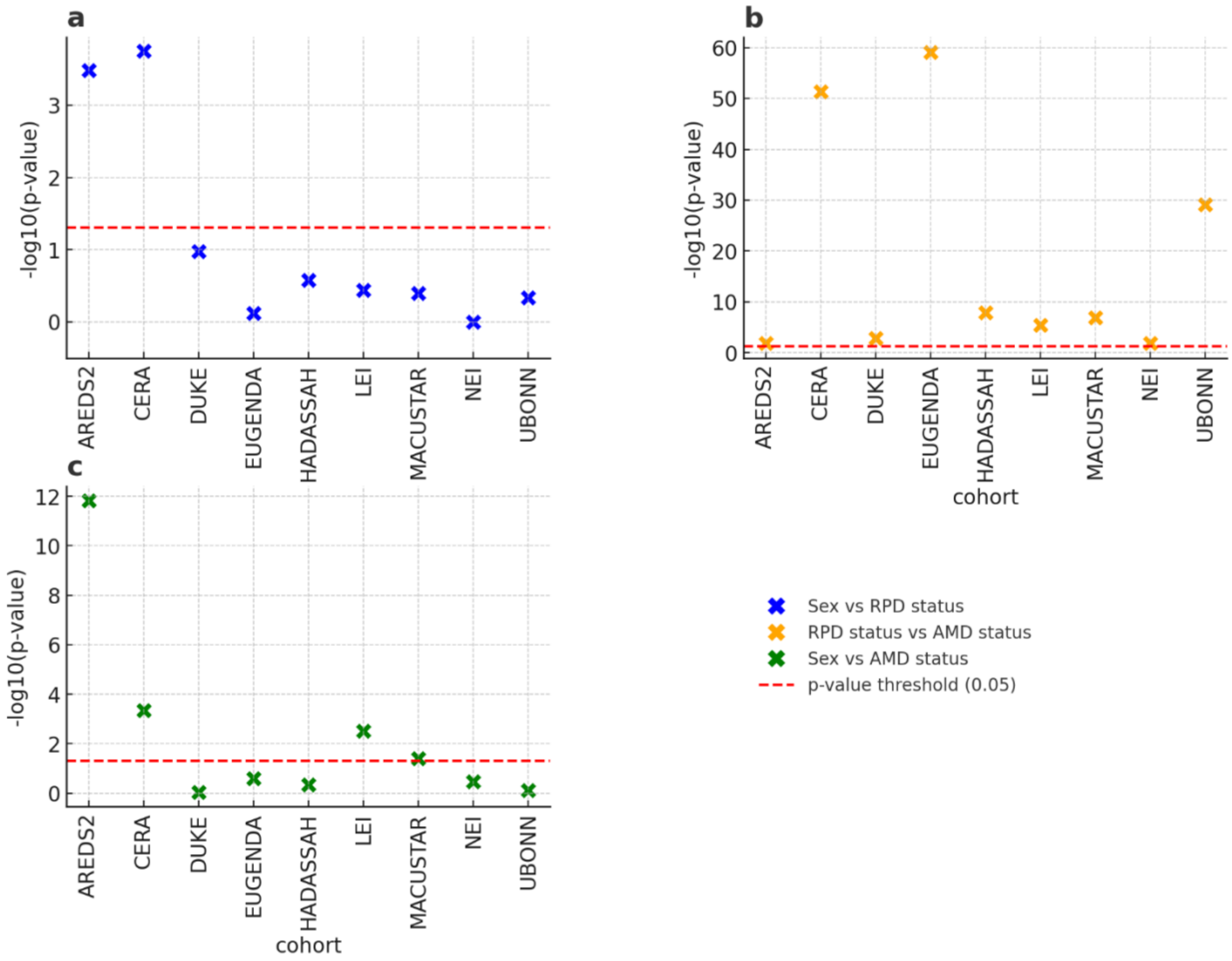


**Supplementary Figure S2. Statistical assessment of demographics and phenotype distributions across cohorts.** The chi-square test was performed to assess **a** The association between sex distribution and RPD status within each cohort; **b** The association between AMD status and RPD status distribution within each cohort; **c** the association between sex distribution and AMD status within each cohort. The red dashed line across all panels marks the -log10(*P-value*) threshold of 1.3 (corresponding to $p = 0.05$). Points above this line indicate statistically significant associations.

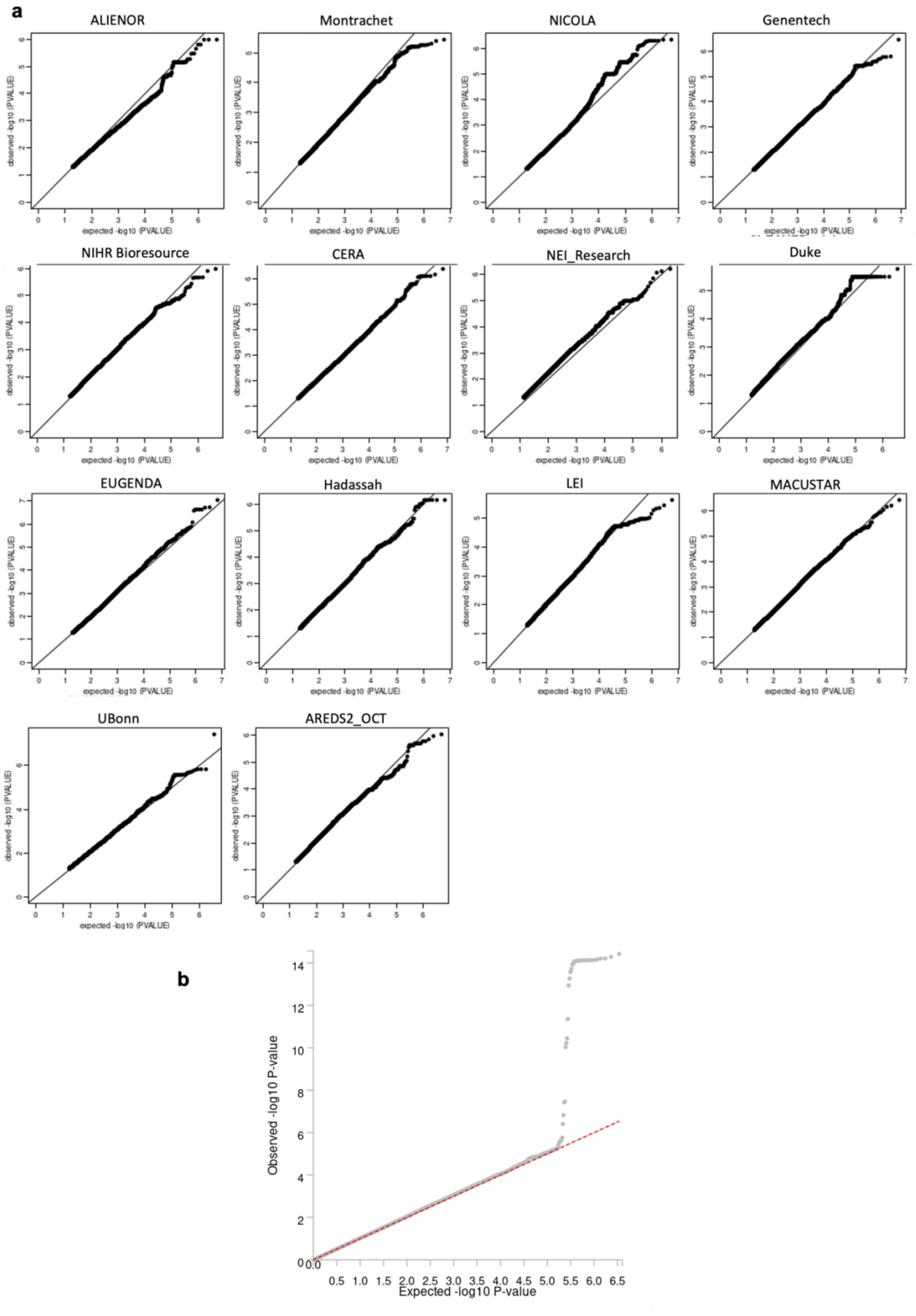

a
ALIENOR
Montrachet
NICOLA
Genentech
NIHR Bioresource
CERA
NEI_Research
Duke
EUGENDA
Hadassah
LEI
MACUSTAR
UBonn
AREDS2_OCT
observed -log10 (PVALUE)
expected -log10 (PVALUE)
b
Observed -log10 P-value
Expected -log10 P-value

**Supplementary Figure S3**: **Quantile-Quantile plots (Q-Q plots) of GWAS summary statistics of AMD+/RPD+ versus AMD+/RPD-. a** Q-Q of GWAS meta-analysis; **b** Q-Q of GWAS of the 14 individual cohorts. Each plot represents a different cohort, with the cohort names indicated in the labels above each plot. The Q-Q plots display the observed p-values versus expected p-values which are the log-transformed p-values, under the null hypothesis have a uniform distribution, indicating that the distribution of test statistics across the cohorts for the genetic association studies follows the expected distribution. This is also formally summarized with the lambda inflation measures (Supplementary Table S4). Note that some cohorts are quite small.

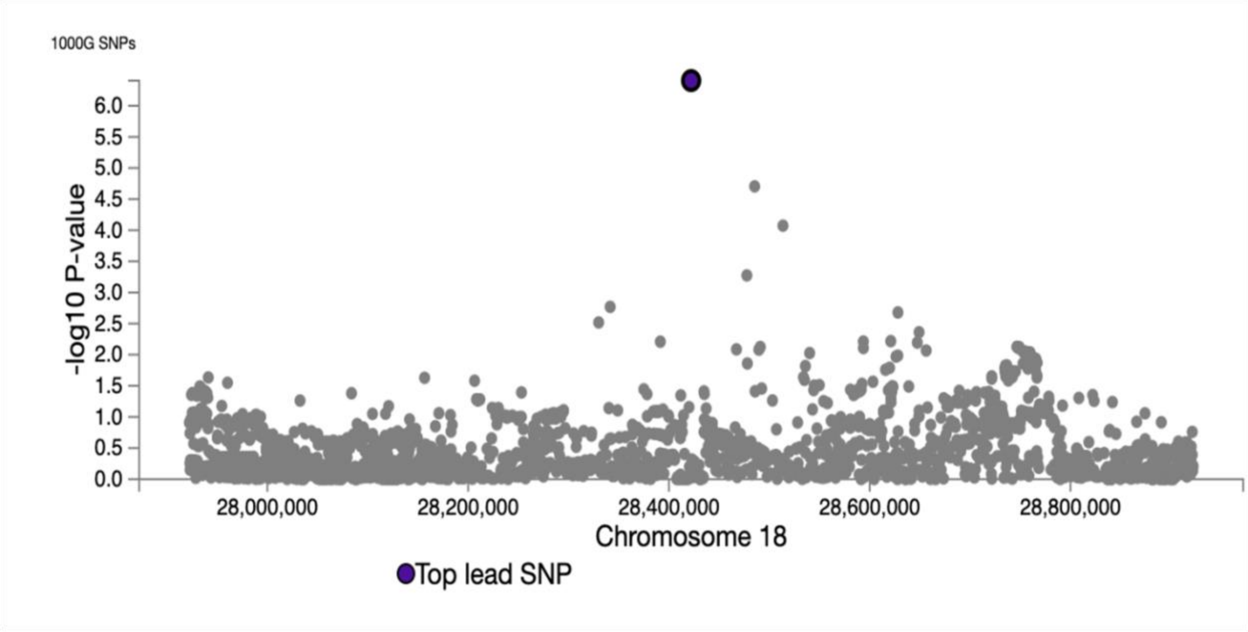


**Supplementary Figure S4.** Regional plot of one SNP on chromosome 18 (SNP rs76361335) identified as associated with RPD at a suggestive significant level (p-value = $1 \times 10^{-6}$). No other SNPs (grey dots) were identified in high linkage disequilibrium (LD) with this SNP, according to the reference genome (1000G Phase 3 European ancestry) using the hg19 genomic build.

**a**

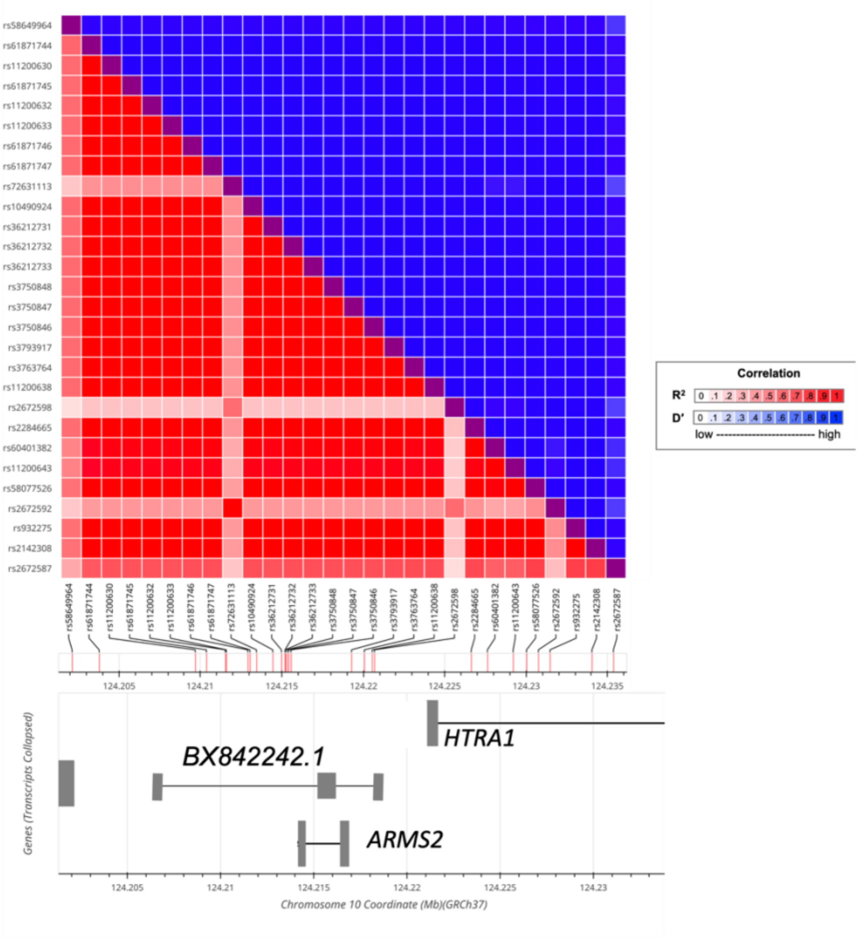
Correlation
R²
D'
low ---------------------------- high
HTRA1
BX842242.1
ARMS2
Genes (Transcripts Collapsed)
Chromosome 10 Coordinate (Mb)(GRCh37)

**b**

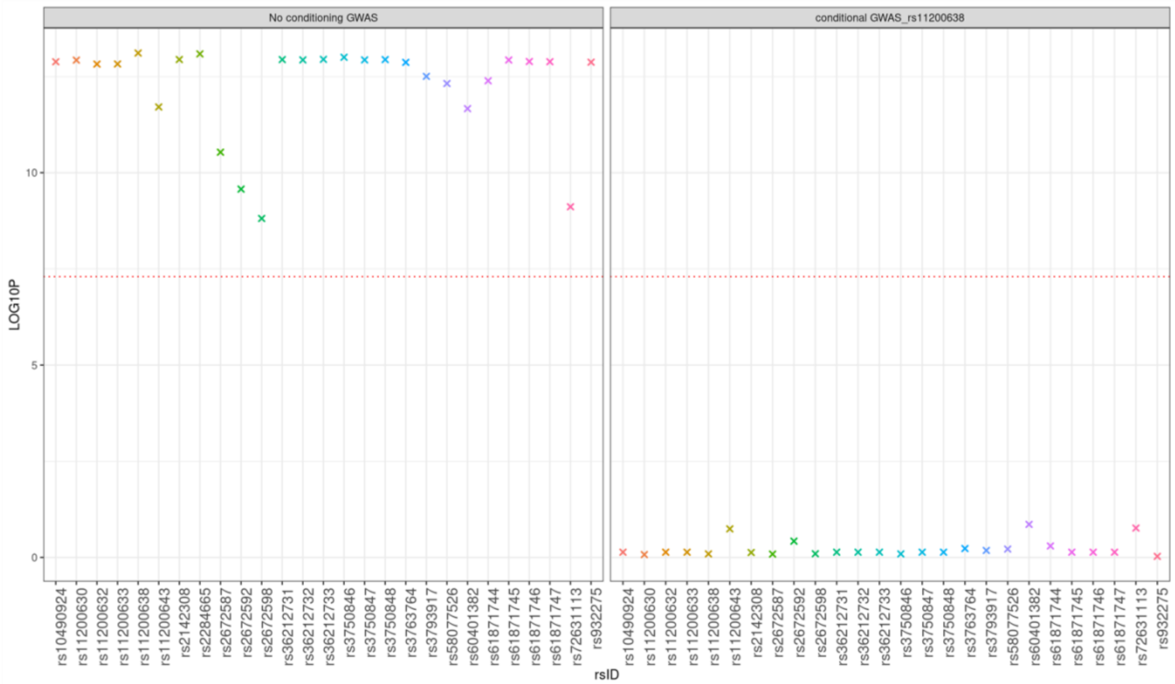
No conditioning GWAS
conditional GWAS_rs11200638
LOG10P
rsID

**Supplementary Figure S5: Variants within RPD risk region. a** Linkage Disequilibrium (LD) matrix and gene annotation in the RPD risk region. Heatmap of pairwise LD measured by both $r^2$ (bottom triangle) and D' (top triangle) between SNPs in the region on chromosome 10. The LD value is indicated by red and blue squares with dark red representing the correlation among these SNPs. *ARMS2*, *HTRA1*, and the long non-coding RNA *BX842242.1* are located within the RPD risk region. SNPs selected for conditional GWAS analysis are marked with asterisks.**; b** Point plot depicting local chr 10 risk locus SNP association for unconditional and conditional GWAS. The left panel depicts GWAS performed without any conditioning, right panel conditional GWAS with RPD risk lead SNP (rs11200638). The X-axis represents the rsIDs, while the Y-axis shows the -log10 p-values. Genome-wide statistical significance threshold is indicated by a red dashed line. Only the unconditional GWAS shows any SNPs in the critical region achieving genome-wide significance.

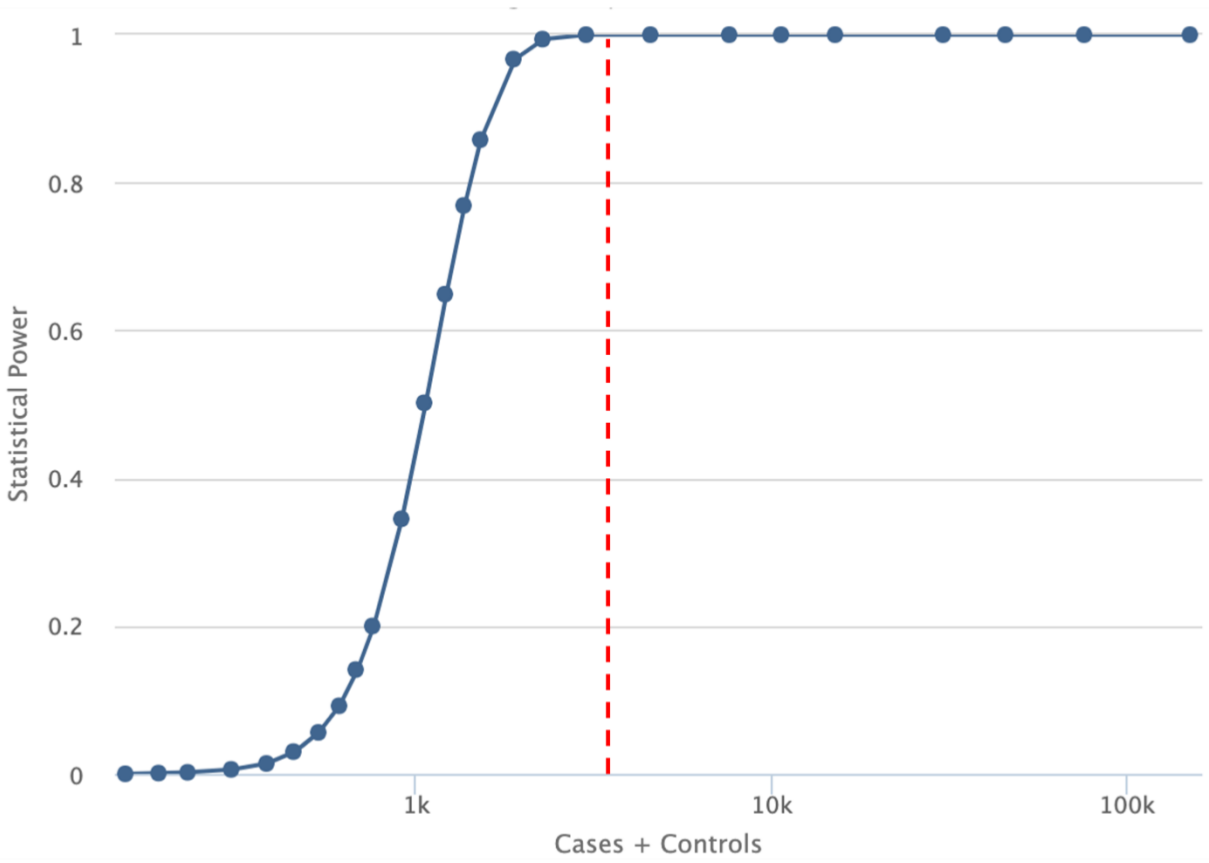


**Supplementary Figure S6: Power calculation to detect genome-wide significant signal on chromosome 1 in GWAS1.** Genetic Association Study Power Calculator (1) was used to compute the power of our RPD-GWAS in detecting chromosome 1 signals. In our testing framework, the top significant SNP on chromosome 1 (rs800292) from a recent AMD-GWAS (2) with beta=-0.7 and MAF=0.23 would be detected in our RPD-GWAS (N= 6,346, indicated by the dashed red line, where Cases/Controls ratio = 0.518) with 100% power at the genome-wide significance level (p-value $< 5 \times 10^{-8}$).

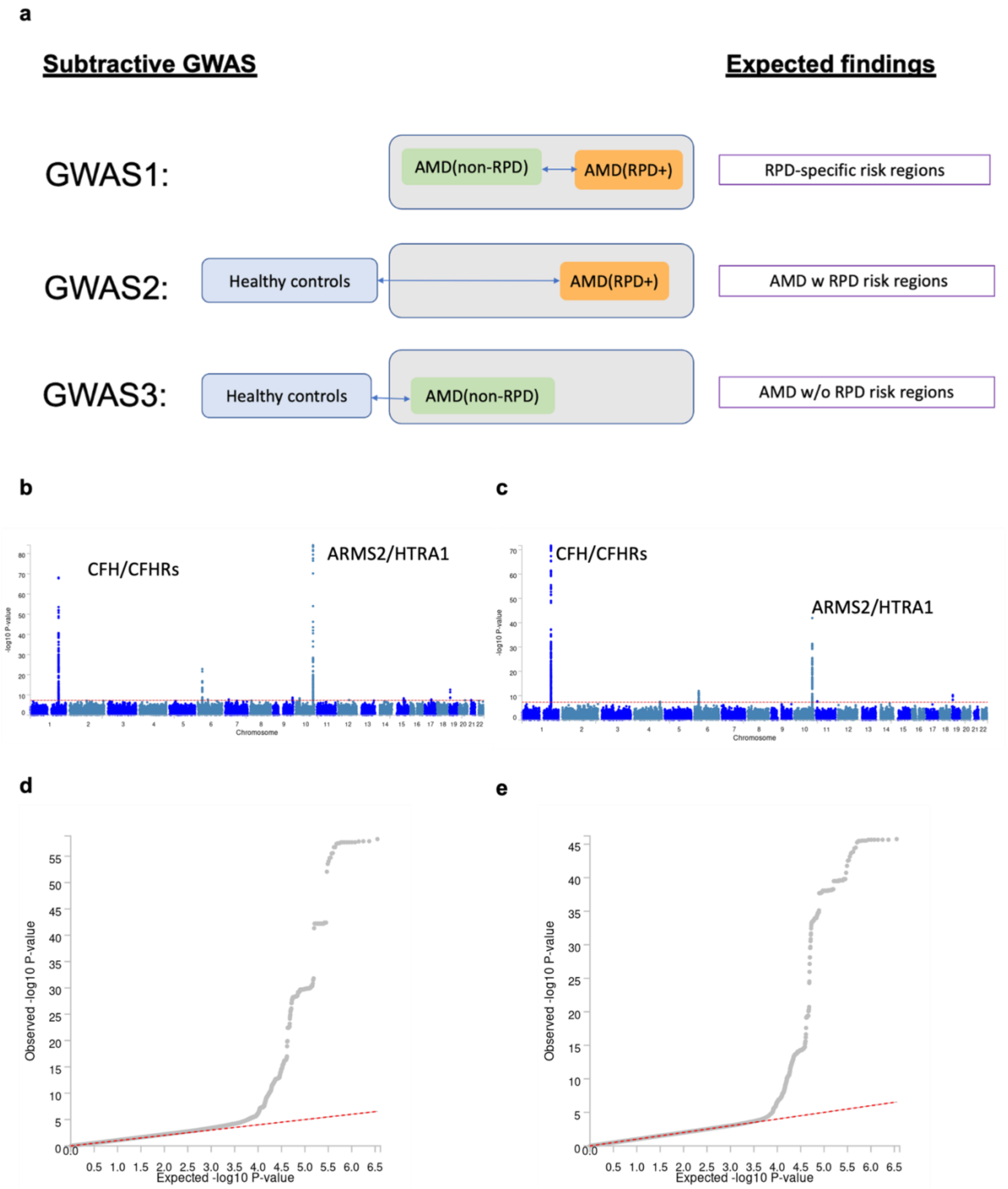


**Supplementary Figure S7: Subtractive GWAS analyses. a** Study design illustrating the comparisons to investigate **(i)** The primary GWAS that compares AMD+/RPD+ individuals to AMD+/RPD- (‘GWAS1’), **(ii)** AMD/+RPD versus controls (‘GWAS2’), and **(iii)** AMD+/RPD- versus controls (‘GWAS3’); **b & c** Manhattan plots showing –log10(P) values across the genome for GWAS2 and GWAS3, respectively; **d & e** Quantile-Quantile plots (Q-Q plots) of GWAS2 and GWAS3, respectively.

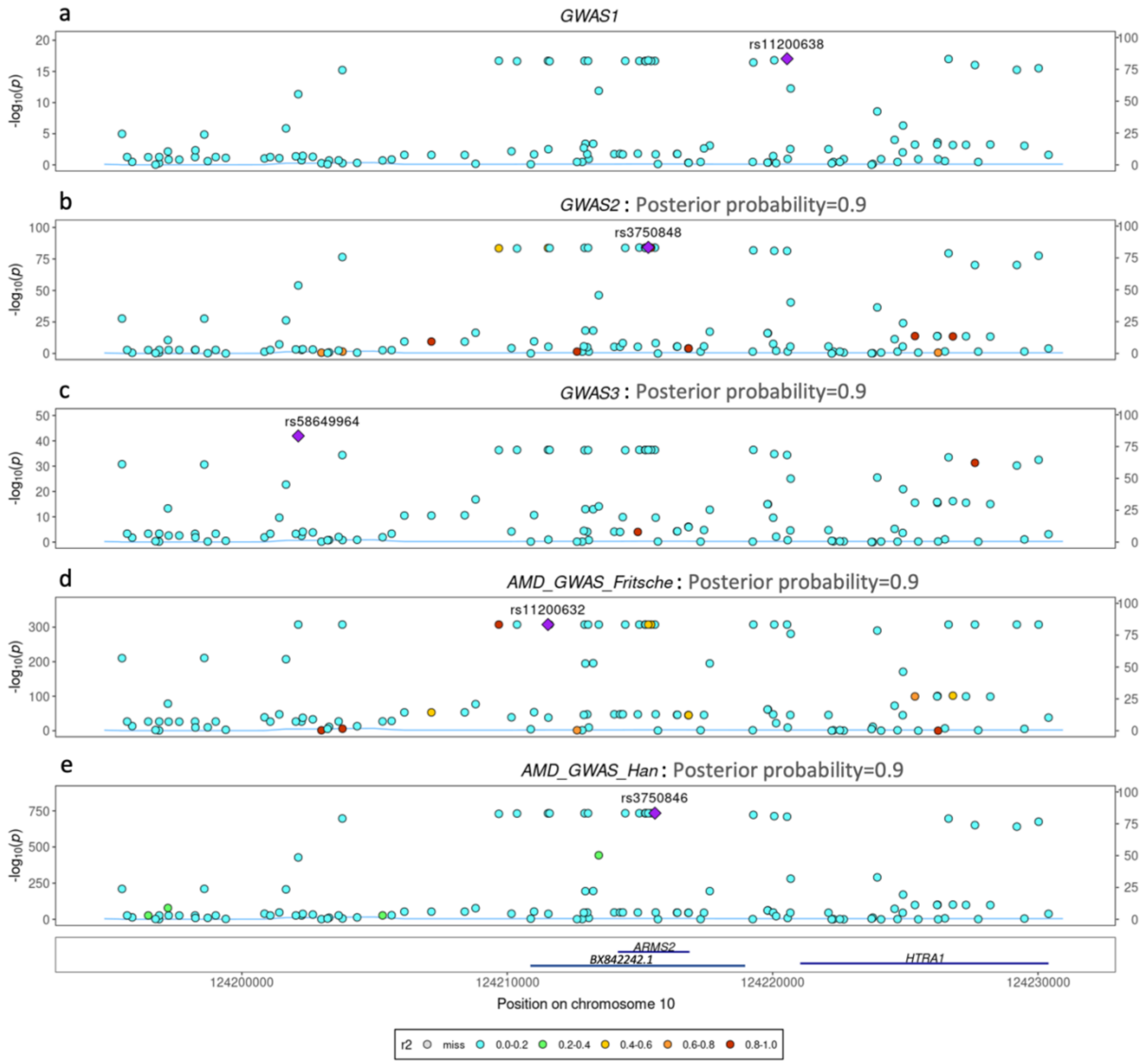


**Supplementary Figure S8:** LocusZoom plots illustrating genetic colocalization analysis, using *coloc* tool version 5.2.2 (3), between RPD Risk region on *ARMS2/HTRA1* locus **(a)** compared to GWAS2 **(b)**, GWAS3 **(c)**, AMD-GWAS study by Fritsche *et al* **(d)**, and AMD-GWAS study by Han *et al* **(e)**. The posterior probability values represent the probability of shared variants among GWASs with GWAS1.

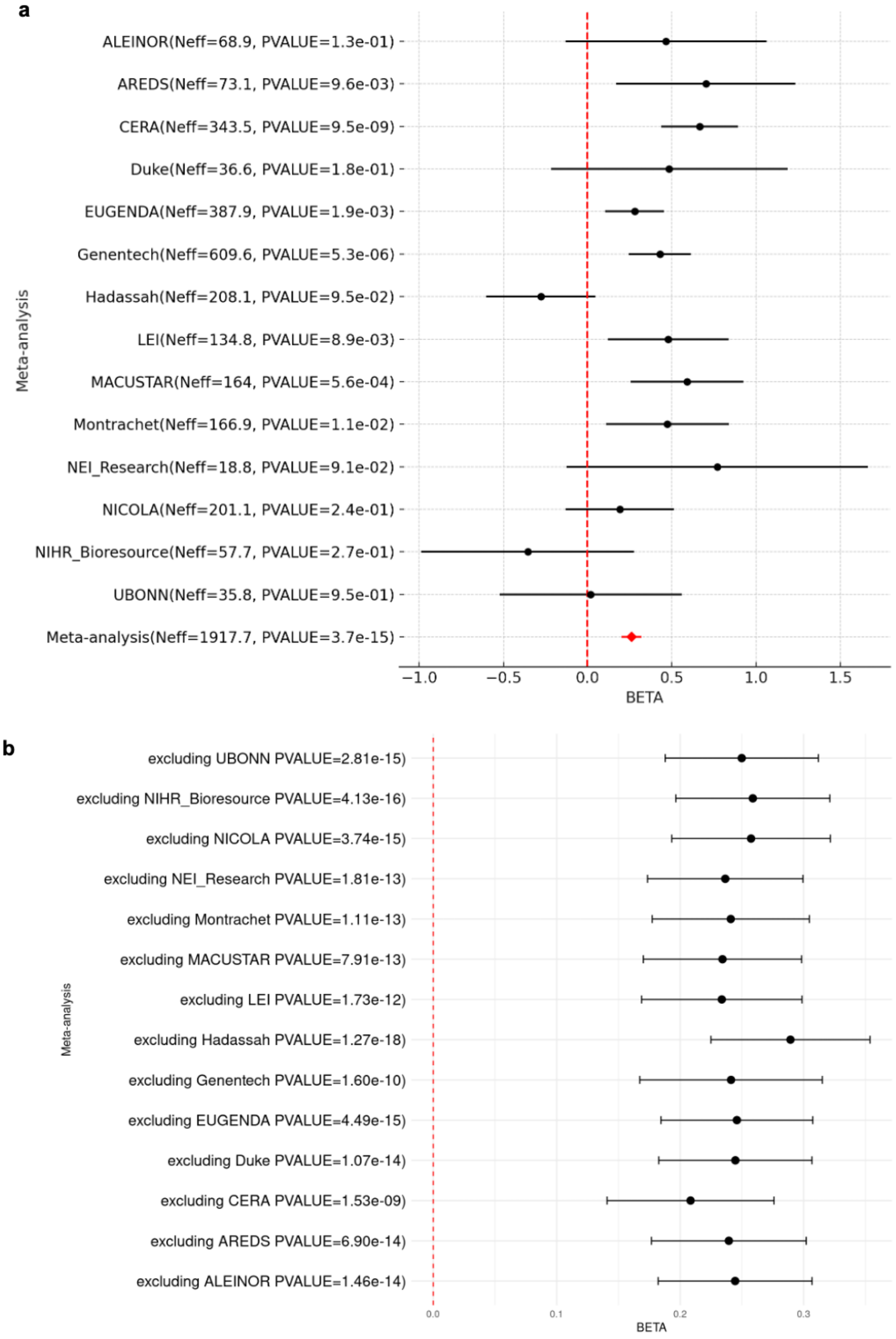
a
ALEINOR(Neff=68.9, PVALUE=1.3e-01)
AREDS(Neff=73.1, PVALUE=9.6e-03)
CERA(Neff=343.5, PVALUE=9.5e-09)
Duke(Neff=36.6, PVALUE=1.8e-01)
EUGENDA(Neff=387.9, PVALUE=1.9e-03)
Genentech(Neff=609.6, PVALUE=5.3e-06)
Hadassah(Neff=208.1, PVALUE=9.5e-02)
LEI(Neff=134.8, PVALUE=8.9e-03)
MACUSTAR(Neff=164, PVALUE=5.6e-04)
Montrachet(Neff=166.9, PVALUE=1.1e-02)
NEI_Research(Neff=18.8, PVALUE=9.1e-02)
NICOLA(Neff=201.1, PVALUE=2.4e-01)
NIHR_Bioresource(Neff=57.7, PVALUE=2.7e-01)
UBONN(Neff=35.8, PVALUE=9.5e-01)
Meta-analysis(Neff=1917.7, PVALUE=3.7e-15)
Meta-analysis
−1.0
−0.5
0.0
0.5
1.0
1.5
BETA
b
excluding UBONN PVALUE=2.81e-15)
excluding NIHR_Bioresource PVALUE=4.13e-16)
excluding NICOLA PVALUE=3.74e-15)
excluding NEI_Research PVALUE=1.81e-13)
excluding Montrachet PVALUE=1.11e-13)
excluding MACUSTAR PVALUE=7.91e-13)
excluding LEI PVALUE=1.73e-12)
excluding Hadassah PVALUE=1.27e-18)
excluding Genentech PVALUE=1.60e-10)
excluding EUGENDA PVALUE=4.49e-15)
excluding Duke PVALUE=1.07e-14)
excluding CERA PVALUE=1.53e-09)
excluding AREDS PVALUE=6.90e-14)
excluding ALEINOR PVALUE=1.46e-14)
Meta-analysis
0.0
0.1
0.2
0.3
BETA

**Supplementary Figure S9**: **Cohort-specific statistical info of RPD lead SNP in GWAS meta-analysis. a** Cohort-specific significance level of the top lead RPD risk SNP 10:124220544:G:A (rs11200638) in the summary statistics of each cohort; **b** Effect size (BETA) and p-values for this SNP in the meta-analyses of cohorts when we exclude one cohort at a time. The red dashed line indicates the effect size value of zero.

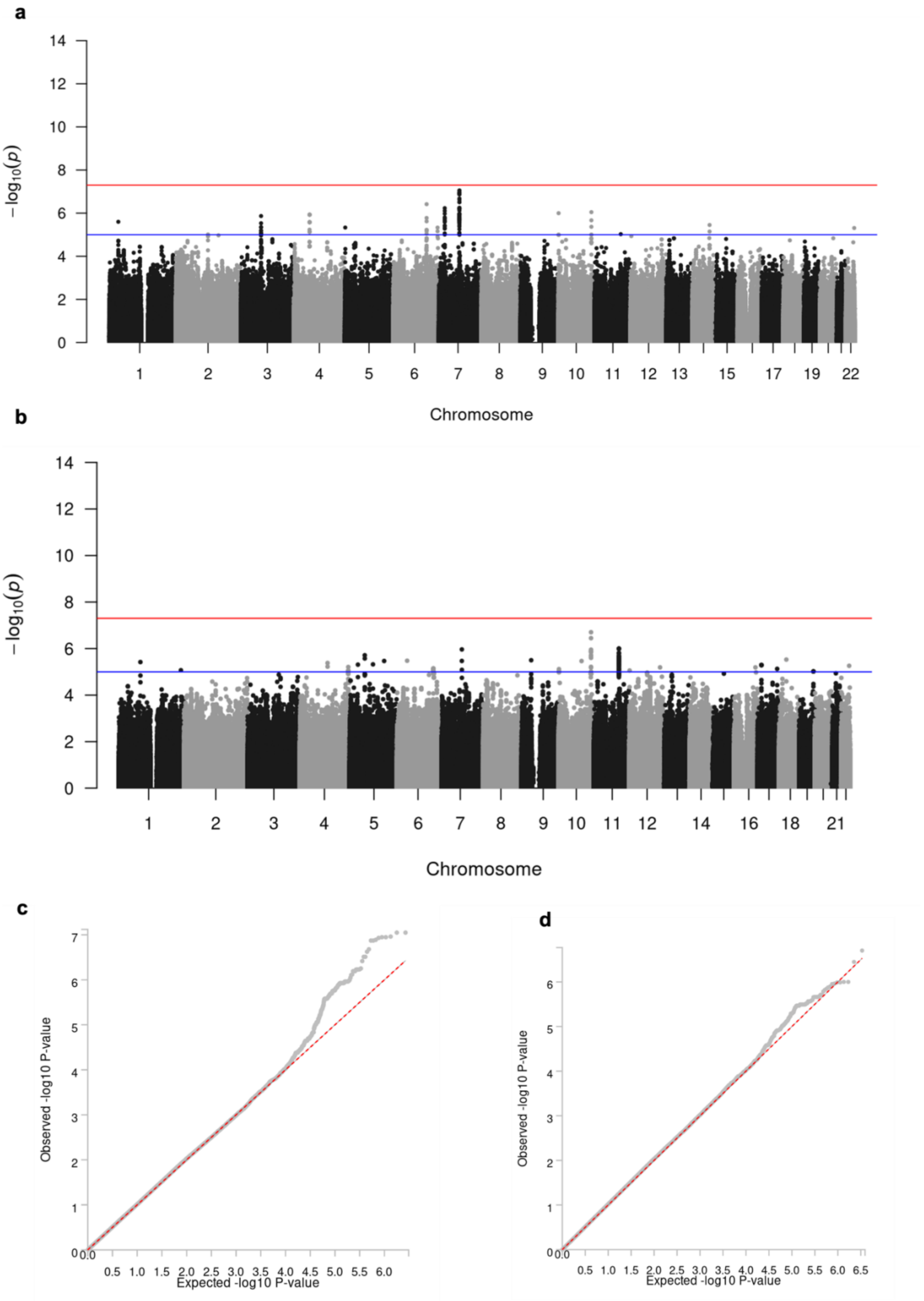

a
b
c
d
−log10(p)
Chromosome
Observed -log10 P-value
Expected -log10 P-value

**Supplementary Figure S10: Manhattan plots for non-additive models of genome-wide association study of RPD. a** The recessive GWAS analysis; **b** the dominant GWAS analysis. The red horizontal line indicates the genome-wide significance threshold (p-value $5x10^{-8}$), and the blue horizontal line represents the suggestive significance level (p-value $1x10^{-6}$); **c** & **d** show Quantile-Quantile plots (Q-Q plots) of recessive and dominant GWAS results, respectively.

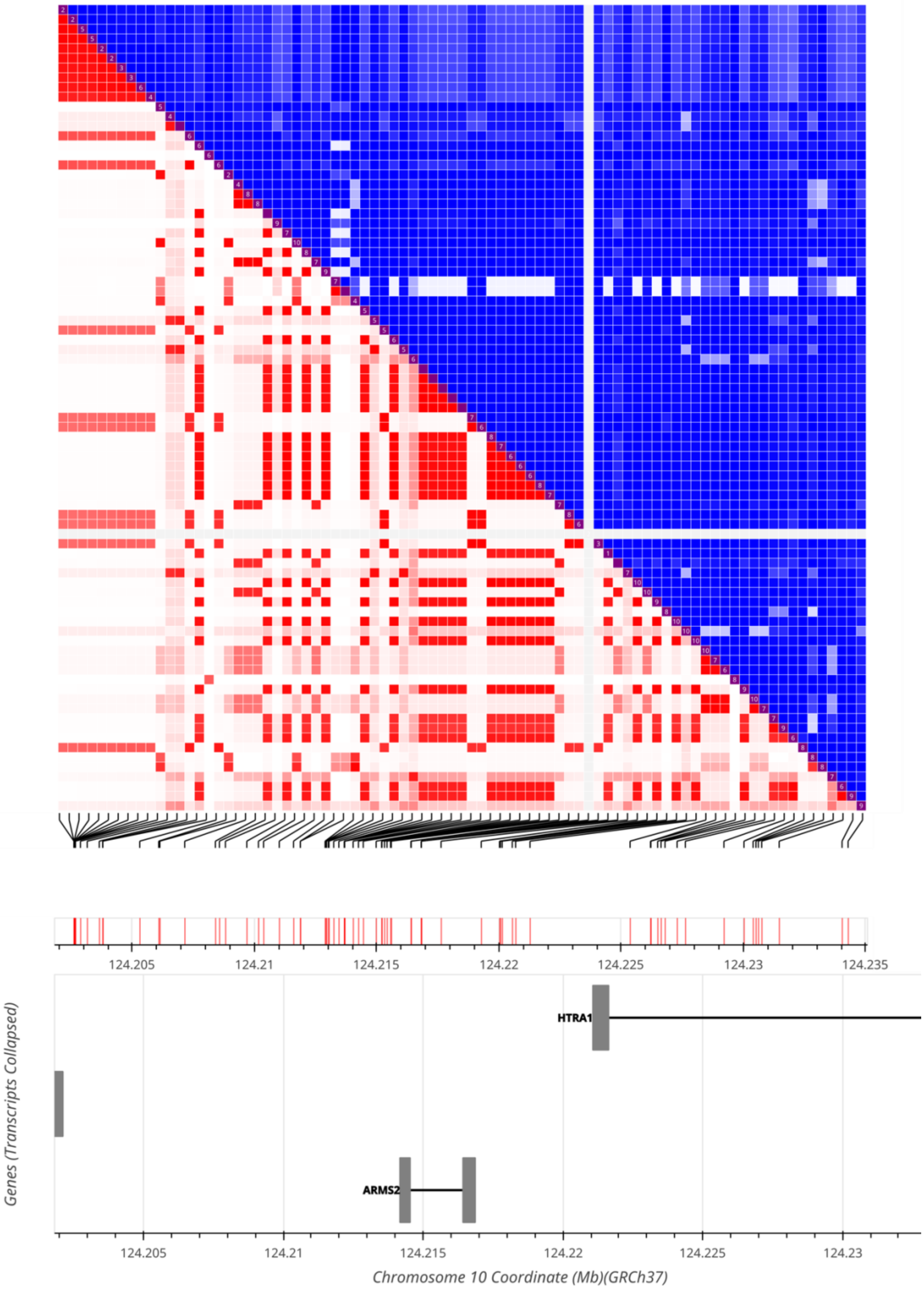

124.205
124.21
124.215
124.22
124.225
124.23
124.235
HTRA1
ARMS2
Genes (Transcripts Collapsed)
Chromosome 10 Coordinate (Mb)(GRCh37)

**Supplementary Figure S11.** The LD matrix of additional SNPs located within genome-wide significant RPD risk region, identified by the WGS method with $r^2$ LD scores.

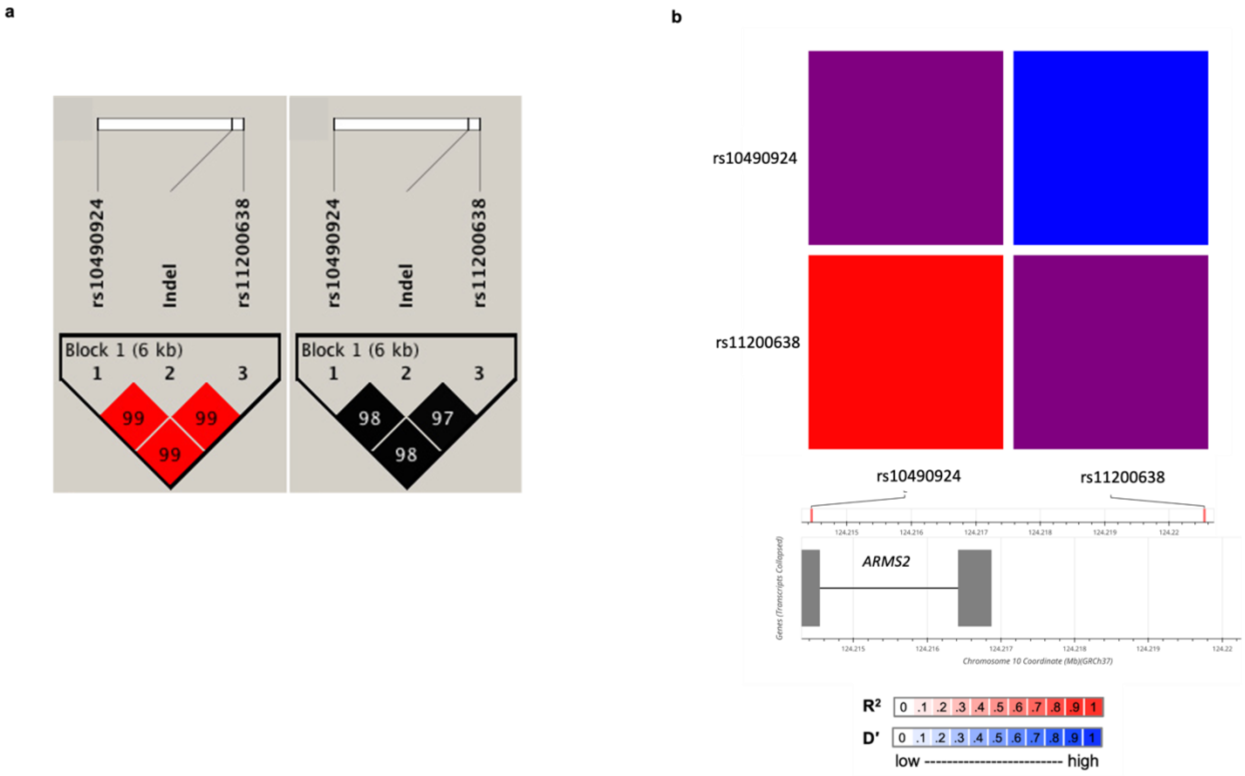


**Supplementary Figure S12: Correlation of RPD-risk lead SNP, proxy SNPs and indel. a** Linkage disequilibrium plots showing the rs10490924, indel and rs11200638 variants in an Australian cohort. The D' (left) and $r^2$ (right) values between SNPs are indicated inside the quadrants for cohorts, figure **a** adapted from Kaur et al., 2013 (4). **b** Heatmap matrix of linkage disequilibrium showing the RPD-risk lead SNP (rs11200638) and rs10490924 in 1000 Genomes Project population groups (n = 2,504) generated by the LDmatrix function of R package "LDlinkR" (5). The Indel is not in the 1000G reference panel.

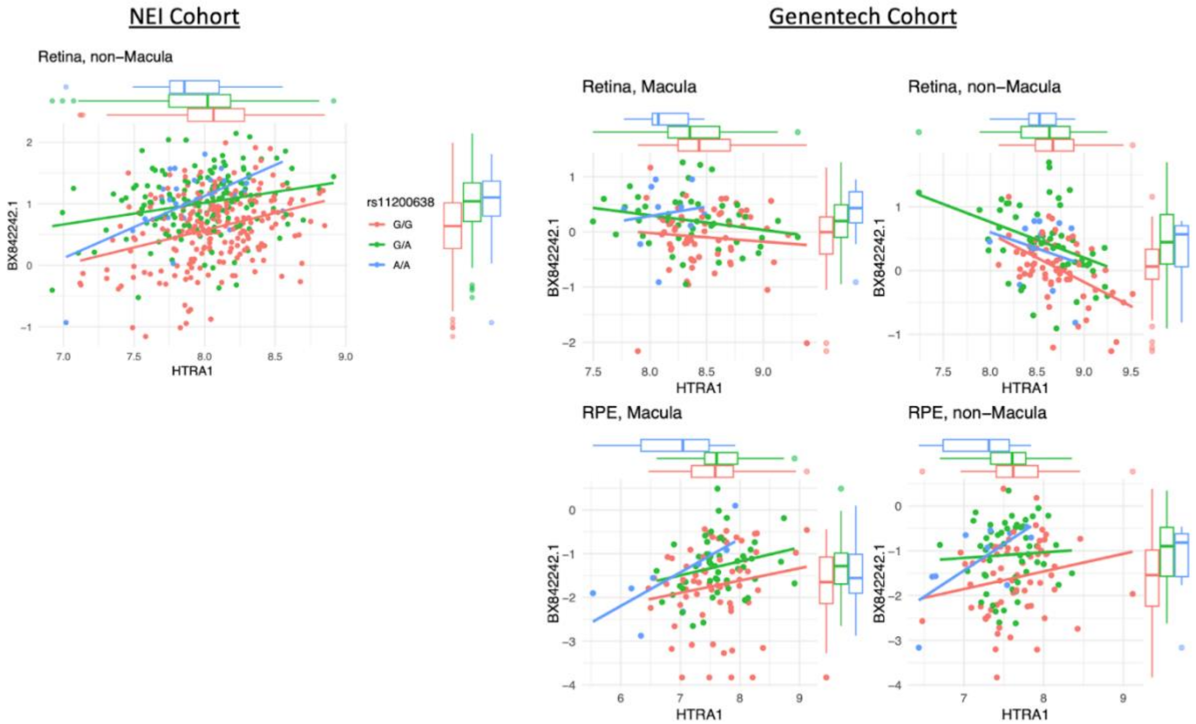


**Supplementary Figure S13.** Expression of HTRA1 RNA compared to *BX842242.1* across retinal tissues from two independent cohorts. Data are colored by the rs11200638 genotype. Marginal boxplots recapitulate eQTL results from Fig. 4. Note that expression correlation is variable across studies and within tissues.

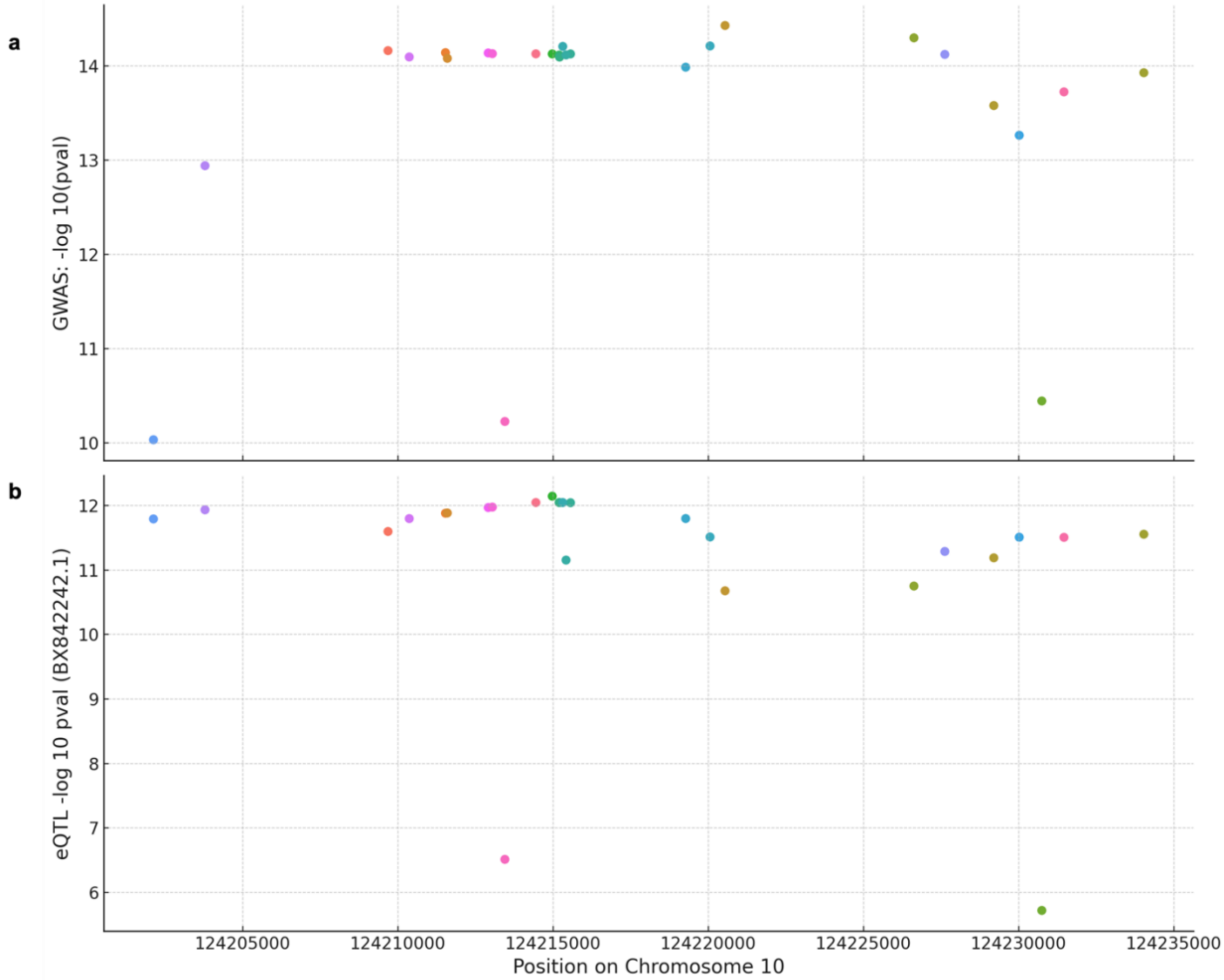


**Supplementary Figure S14: eQTL-GWAS pairs. a** RPD-GWAS results for SNPs (each dot) that are genome-wide significant. **b** eQTL results. The Y-axis present the -log10 (*pval*) for GWAS results and -log10(nominal *pval*) for the *BX842242.1* gene in Strunz et al. (6) Different colours indicate individual SNPs. The x-axis shows the chromosomal positions.

a b

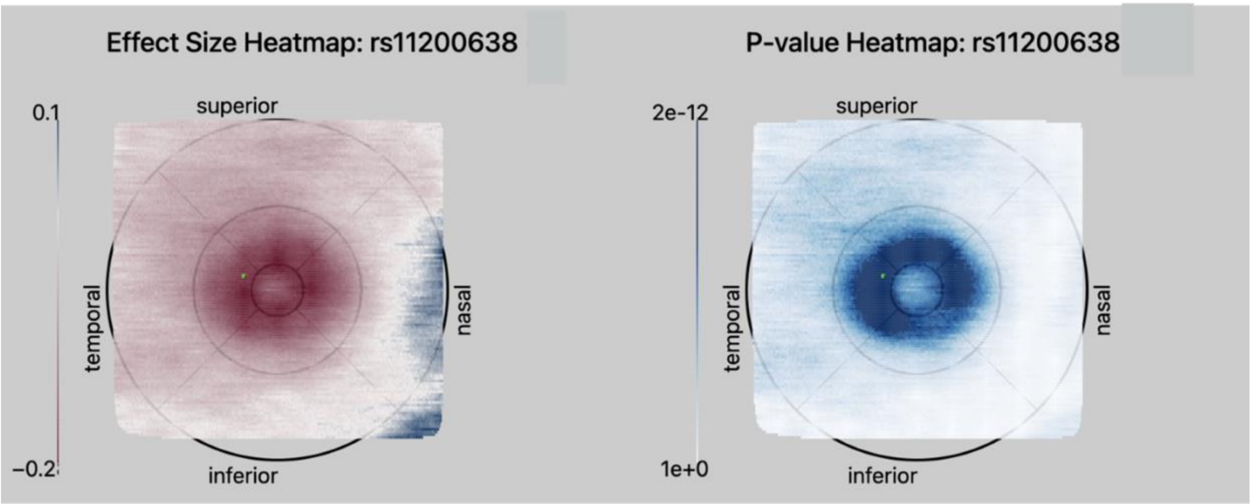


**Supplementary Figure S15: The ETDRS grids of central 6mm region in pixel-wise retinal thickness GWAS results for RPD risk lead SNP (rs11200638). a** Effect size on retinal thickness across pixels for the SNP rs11200638. The color gradient depicts the range of effect sizes, positive values in blue indicate an increase in retinal thickness, while negative values in red represent a decrease in retinal thickness; **b** P-values of association analysis across pixels for SNP rs11200638. The blue color gradient represents levels of statistical significance with dark blue indicating higher statistical significance. Figures were generated from queries to the https://retinomics.org/ webpage (7). Three rings indicate fovea, parafovea and perifovea on ETDRS grid, 6000 microns (μm).

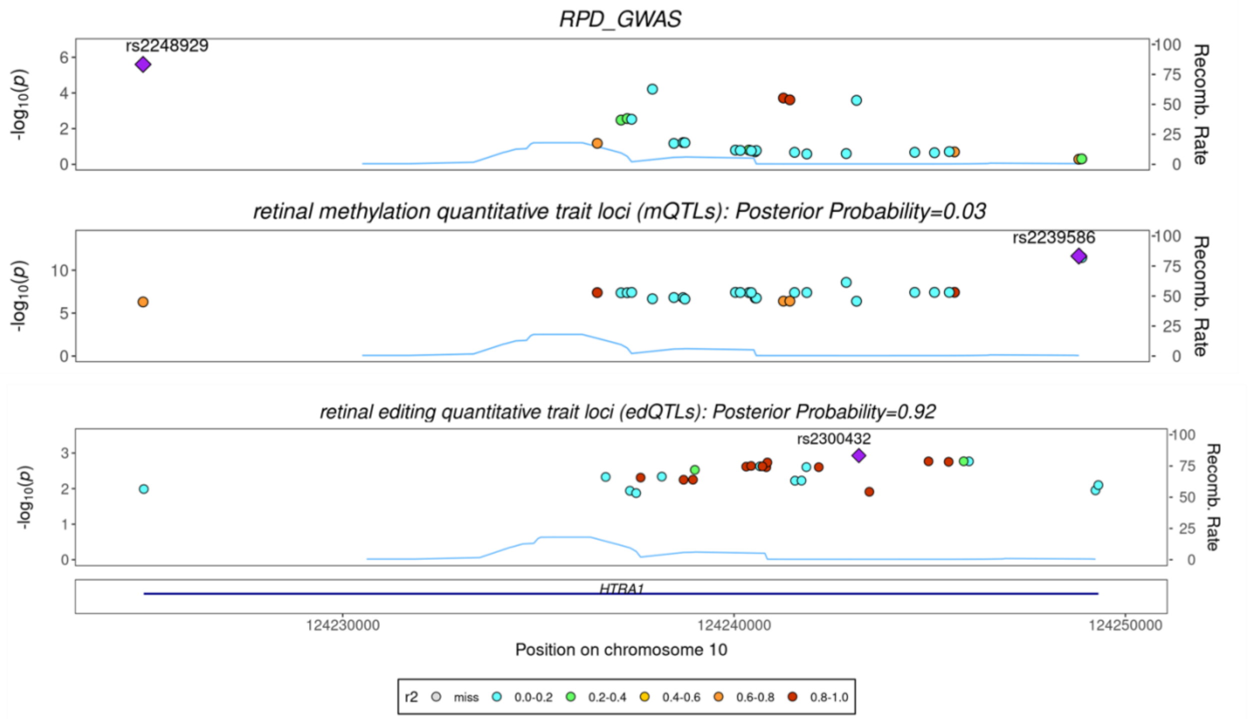


**Supplementary Figure S16:** Colocalization and posterior probability of the chr10 RPD risk locus with retinal methylation quantitative trait loci (mQTL) and retinal editing quantitative trait loci (edQTLs). Each plot includes a recombination rate track (blue line) and a linkage disequilibrium (LD) color scale indicating the $r^2$ values of SNPs in relation to RPD-risk lead SNP (rs11200638). The x-axis represents the position on chromosome 10, and the y-axis represents the -log10(p-value) of association. The blue bars on the x-axis label the genes. lncRNA *BX842242.1* was added manually. The purple diamond represents the most significant SNP in the region for each trait and the dark blue square represents the RPD-risk lead SNP (rs11200638) in each trait.

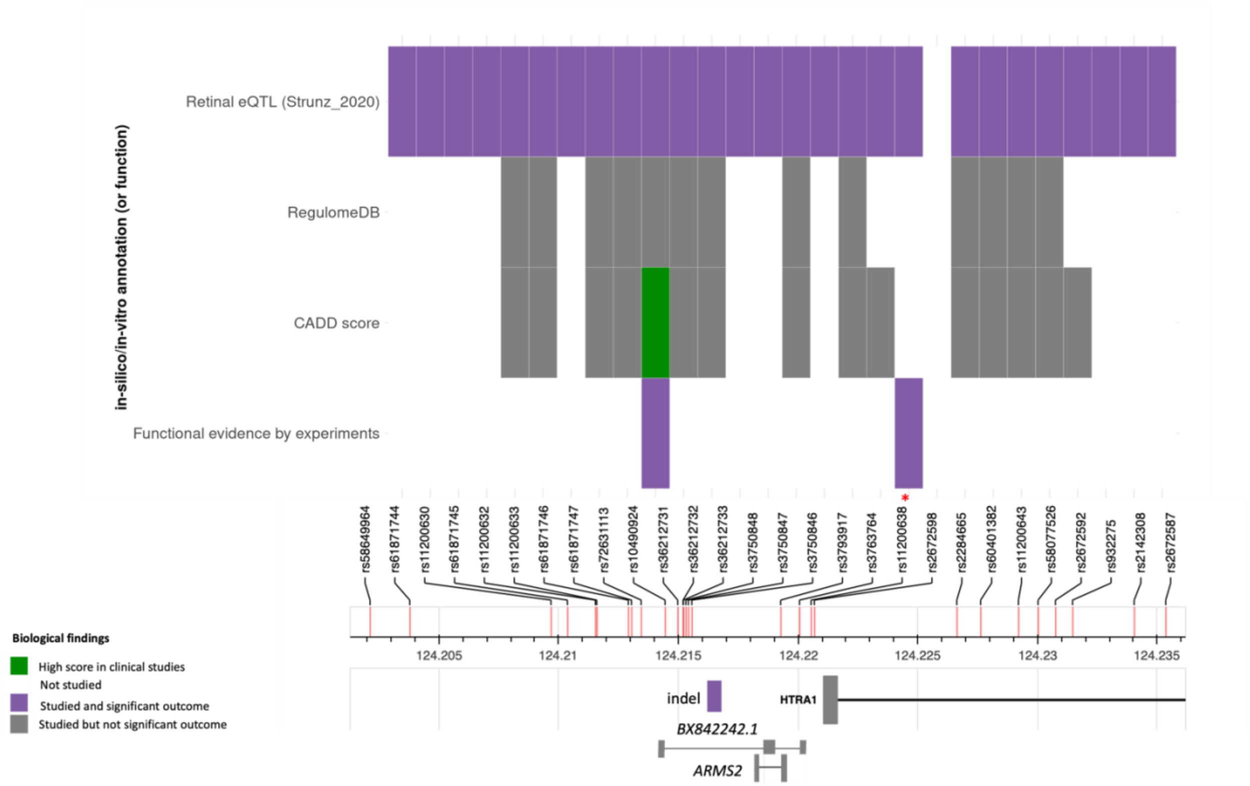


**Supplementary Figure S17: Overview of genome-wide significant RPD-risk associated single nucleotide polymorphisms (SNPs) identified by GWAS and the known functions of variants within ARMS2/HTRA1 locus.** 28 risk variants at the RPD risk locus that are associated with AMD. The variants with functional effects identified or known as eQTLs are indicated in the purple bars. Variants with low RegulomeDB scores (8) (functionally important) are colored in grey. Variants with high CADD scores (9) (clinically important variants) are colored in green. Empty bars represent the SNPs with no data available. RPD-risk lead SNP (rs11200638) is indicated by a red star. The eQTL results for the *BX842242.1* gene in based on Strunz et al. (6). The presumed correlation between rs10490924 and methylation level is based on Oliver et al. (10) (whole blood). The functional effect of the indel (also known as del443ins54) on HTRA1 expression is based on Yang et al. (11) (RPE cells), Iejima et al. (12) (661W, immortalized mouse photoreceptor cell line), Oura et al. (13) (Y79, human retinoblastoma cells) and Pan et al., (14) (661W, HEK-293, COS-7, iPSCs, and whole blood). The correlation of rs11200638 and HTRA1 transcription is based on Yang et al. (15) (lymphocytes and RPE), Chan et al. (16) (retinal cells), Tuo et al. (17) (ocular tissue), Mohamad et al. (18) (whole blood), and Yang et al. (11) (placental tissue).

a

b

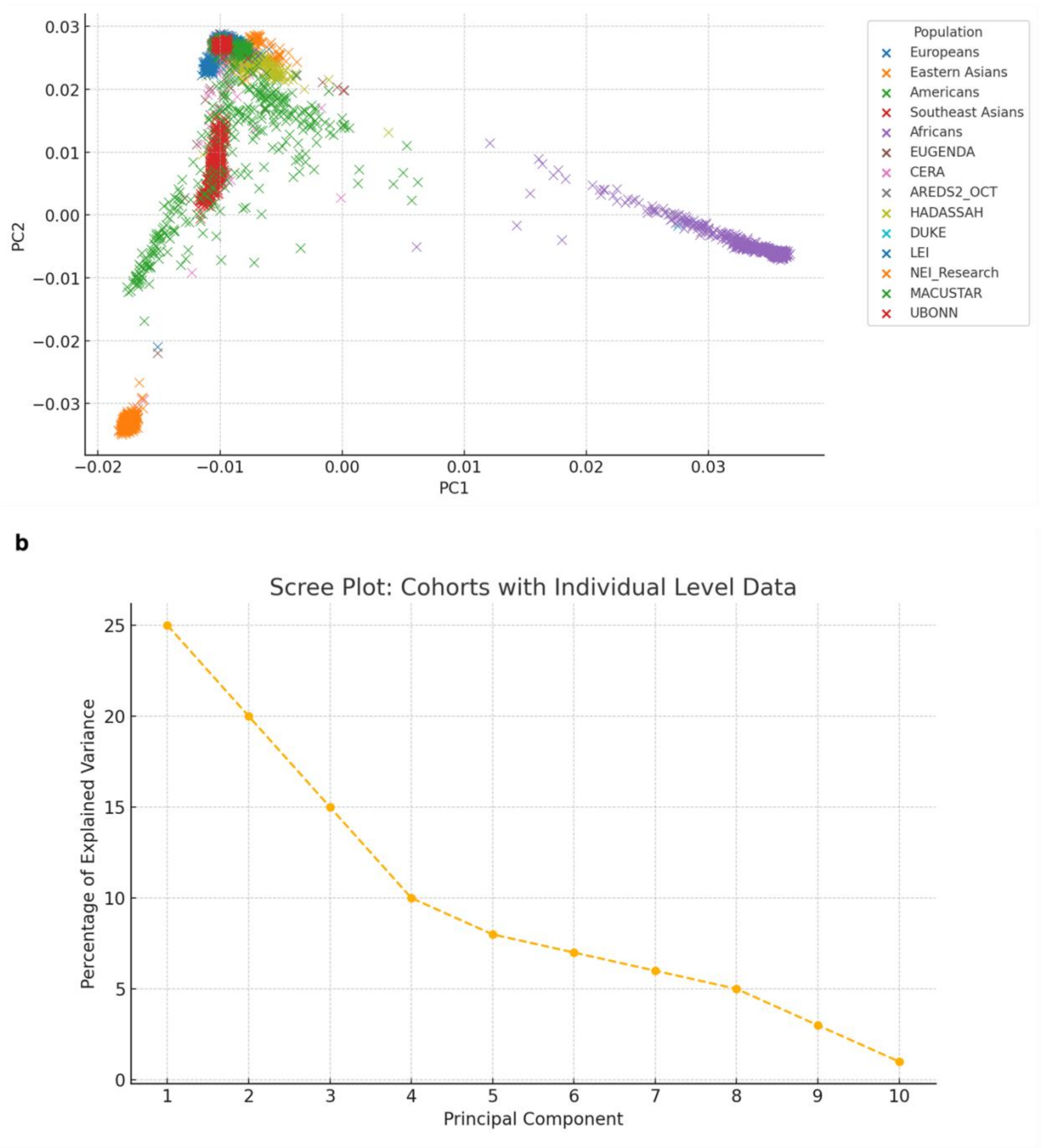


**Supplementary Figure S18**: Principal Component Analysis (PCA) of the nine cohorts with individual-level genotype data. **a** Genetic ancestry distribution of individuals from nine cohorts, overlaid with reference populations (Europeans, Eastern Asians, Americans, Sout East Asians and African) from the 1000 Genomes Project. Each point represents an individual, colored by their population group. Note that the demographic data for the NICOLA cohort is not shown in this figure due to the unavailability of data; **b** Scree plot that depicts the proportion of variance explained by each principal component (PC) in the dataset. The first five PCs capture most of the variability (about 72%), capturing most of the underlying population structure. Note that the demographic data for the NICOLA cohort is not shown in this figure.
